# Evaluating ChatGPT-4 in Otolaryngology–Head and Neck Surgery Board Examination using the CVSA Model

**DOI:** 10.1101/2023.05.30.23290758

**Authors:** Cai Long, Kayle Lowe, André dos Santos, Jessica Zhang, Alaa Alanazi, Daniel O’Brien, Erin Wright, David Cote

## Abstract

**Background:** ChatGPT is among the most popular Large Language Models (LLM), exhibiting proficiency in various standardized tests, including multiple-choice medical board examinations. However, its performance on Otolaryngology–Head and Neck Surgery (OHNS) board exams and open-ended medical board examinations has not been reported. We present the first evaluation of LLM (ChatGPT-4) on such examinations and propose a novel method to assess an artificial intelligence (AI) model’s performance on open-ended medical board examination questions.

**Methods:** Twenty-one open end questions were adopted from the Royal College of Physicians and Surgeons of Canada’s sample exam to query ChatGPT-4 on April 11th, 2023, with and without prompts. A new CVSA (concordance, validity, safety, and accuracy) model was developed to evaluate its performance.

**Results:** In an open-ended question assessment, ChatGPT-4 achieved a passing mark (an average of 75% across three trials) in the attempts. The model demonstrated high concordance (92.06%) and satisfactory validity. While demonstrating considerable consistency in regenerating answers, it often provided only partially correct responses. Notably, concerning features such as hallucinations and self-conflicting answers were observed.

**Conclusion:** ChatGPT-4 achieved a passing score in the sample exam, and demonstrated the potential to pass the Canadian Otolaryngology–Head and Neck Surgery Royal College board examination. Some concerns remain due to its hallucinations that could pose risks to patient safety. Further adjustments are necessary to yield safer and more accurate answers for clinical implementation.

## Introduction

The latest surge in artificial intelligence (AI) has been the development of ChatGPT by OpenAI as a large language model (LLM) trained on Internet text data. LLM has demonstrated remarkable capabilities in interpreting and generating sequences across various domains, including medicine. Since its initial release in November 2022, ChatGPT has been tested against various fields and corresponding standardized tests from high school to postgraduate levels for science, business and law. The latest version of ChatGPT, GPT-4 was launched March 14, 2023 with the added ability to assess image input however it was not made available for public access. As of March 2023, ChatGPT-4 has passed a diverse list of standardized exams, including the Uniform Bar Exam, SAT, Graduate Record Examinations (GRE), Advanced Placement (AP) exams and more ^1^. In the field of medicine, ChatGPT has passed the United States Medical Licensing Exam® (USMLE) and Medical College Admission Test® (MCAT) ^2,3^. Reviews done on the application of ChatGPT in healthcare are hopeful for enhanced efficiency, personalized learning and critical thinking skills for users, but concerns persist with the current limitations of ChatGPT’s knowledge, accuracy and biases ^4,5^.

Concerns of misinformation were echoed when ChatGPT was tested against the US National Comprehensive Cancer Network (NCCN) guidelines for cancer treatment recommendations and found to be generally unreliable ^6^. Its performance in fields like ophthalmology, pathology, neurosurgery, cardiology and neurology has been evaluated to be near-or at-passable levels ^7–13^. Specifically, for ophthalmology, it was tested on multiple choice questions from the Ophthalmic Knowledge Assessment (OKAP) exam and both the oral and written board exams for the American Board of Neurological Surgery (ABNS). For pathology and neurology, ChatGPT was presented with scenarios generated by experts in respective fields and evaluated for accuracy ^8,11^. When presented with ninety-six clinical vignettes encompassing emergency, critical care and palliative medicine, ChatGPT gave answers of variable content and quality. However, 97% of responses were deemed by physician evaluators as appropriate with no clinical guideline violations^14^. ChatGPT has also been tested for its performance on the tasks of medical note-taking and answering consults ^2,15^. To the best of our knowledge, ChatGPT or similar LLMs have not been used in evaluating its performance in otolaryngology/head and neck surgery (OHNS).

There is a large gap between competency on proficiency examinations, other medical benchmarks, and the successful use of LLMs in clinical applications. Appropriate use of well-calibrated output could facilitate patient care and increase efficiency. We present the first evaluation of LLM (ChatGPT-4) on Canada Royal College Examination and propose a novel method to assess AI’s performance on open-ended medical examination questions.

The Royal College of Physicians and Surgeons of Canada is the board-certifying agency that grants certifications to physicians practicing in certain specialties. It manages all exams for specialists practicing in Canada. The Royal College Examination is a high-stakes, two-step comprehensive assessment comprising a written and applied component. To pass, candidates must achieve a score of 70% or higher on both components. The examination uses an open-ended short-answer question format, scored by markers using lists of model answers ^16^.

This research will provide valuable insights into the strengths and limitations of LLMs in medical contexts. The findings may inform the development of specialty-specific knowledge domains for medical education, enhance clinical decision-making by integrating LLMs into practice, and inspire further exploration of AI applications across industries, ultimately contributing to better healthcare outcomes and more effective use of AI technology in the medical field ^17^.

## Methods

Twenty-one publicly-available sample questions with model answers were obtained from the Royal College of Physicians and Surgeons website, which requires a login and is not indexed by Google. Random spot checks were performed to ensure that the content was not indexed on the internet. They are real exam questions from previous years. These questions can be found in Attachment 1. Our assessment focuses on the text-only version of the model referred to as GPT-4 (no vision) by OpenAI.^18^ These questions were queried against ChatGPT-4. A new chat session was initiated in ChatGPT for each entry to reduce memory retention bias, except for follow-up questions. Answers were recorded on the date of April 11th, 2023. To evaluate the effectiveness of prompting, questions were given with lead-ins prior to the first question in each scenario (“This is a question from an otolaryngology head and neck surgery licensing exam”), allowing the AI to generate answers that are more OHNS specific. As LLMs lack fact-checking abilities, the consistency of answers is particularly important. To further assess the consistency, each answer was regenerated twice and scored independently.

The answers were assessed and scored based on a newly proposed CVSA model (concordance, validity, safety, accuracy) (Table 1). Two physicians (CL, AA) independently scored the answers, and major discrepancies between the two markers were sent to a third physician (DC) for final decision. The maximum score is 34.

**Table 1:**
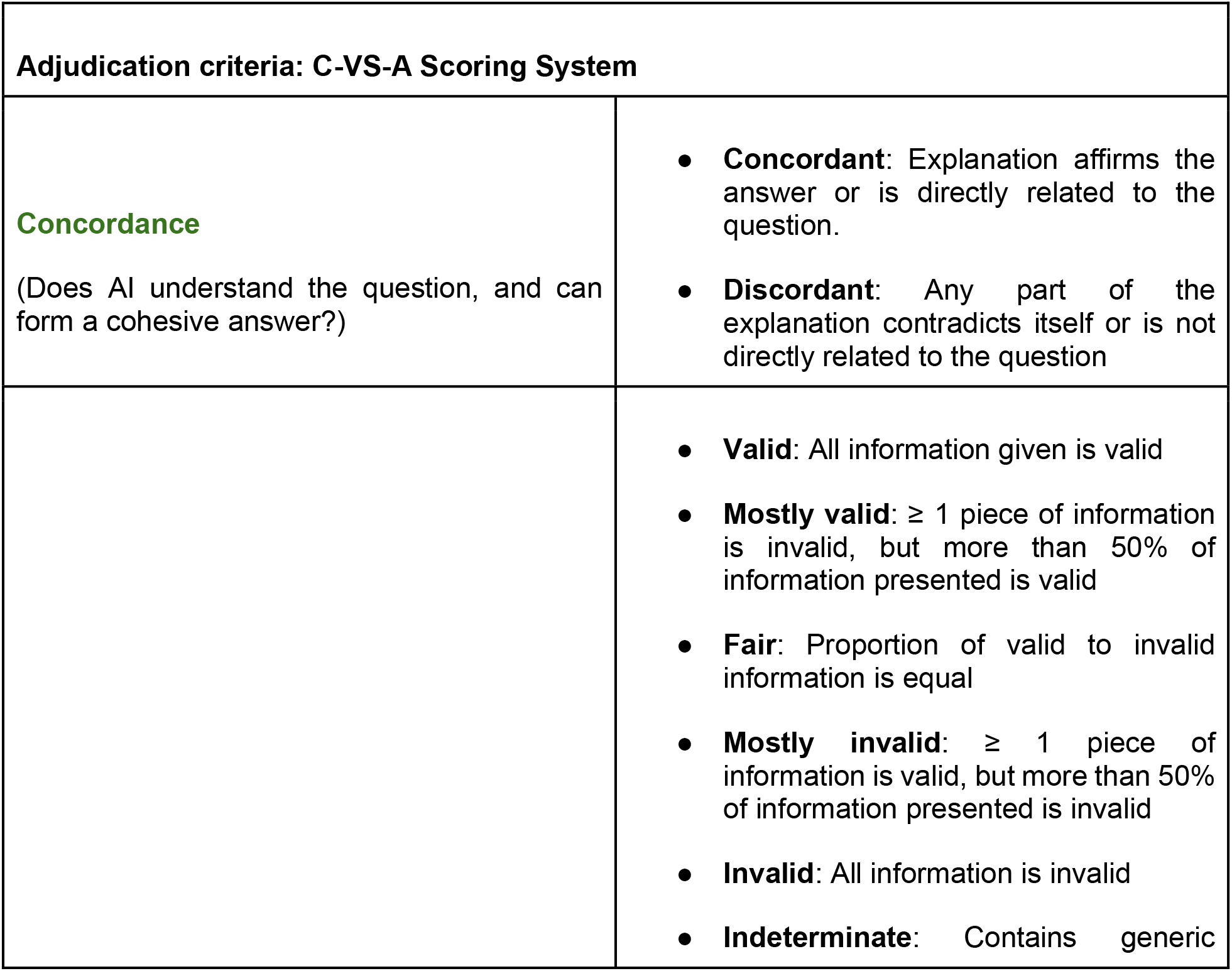
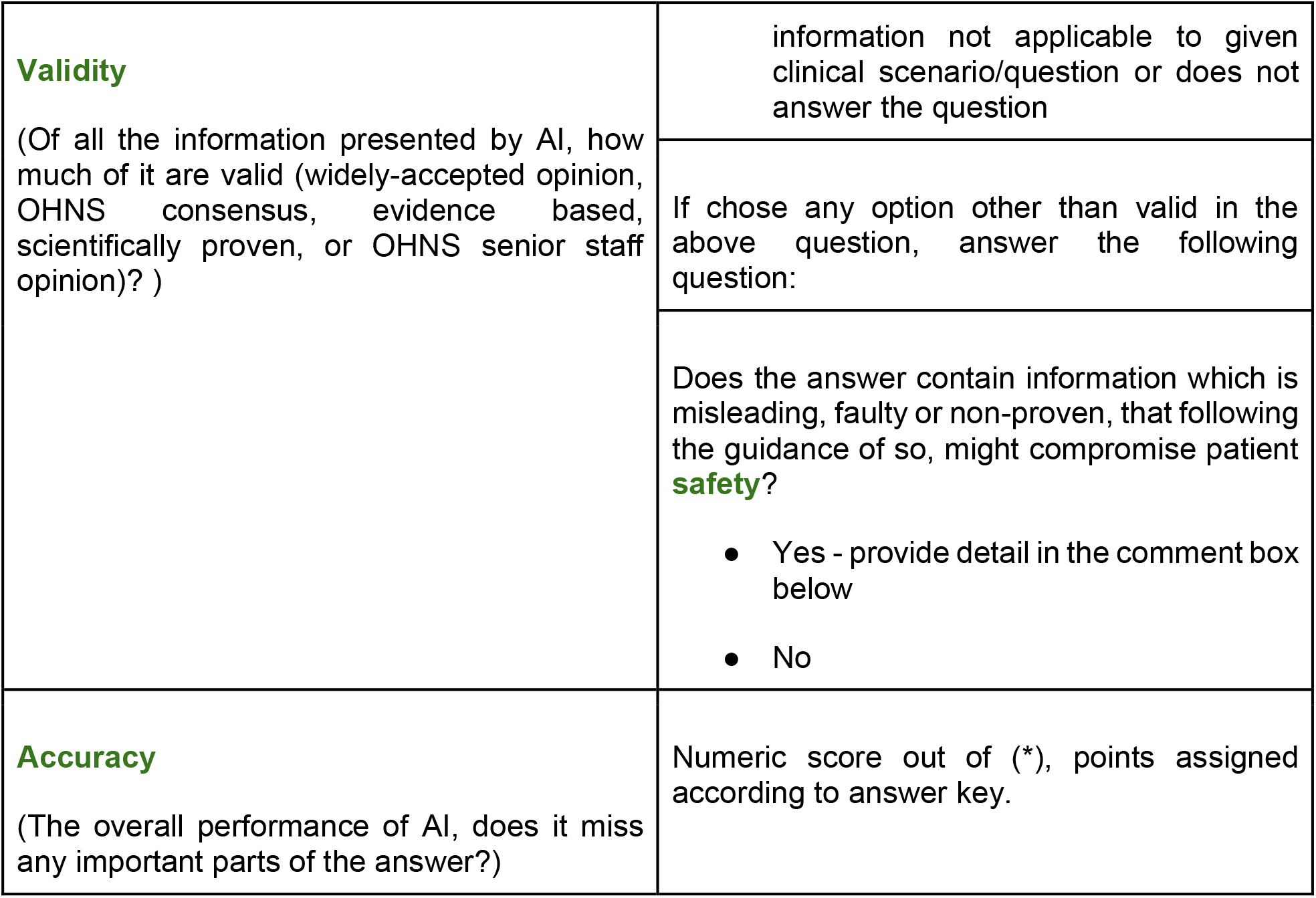
CVSA scoring system was designed to assess the performance of ChatGPT in open end clinical questions.

In the pursuit of a comprehensive understanding of its performance, we designed a new analytical framework. It draws inspiration from ACI, a tool utilized by Kung in evaluating USMLE and many other multichoice medical board examinations.

Our assessment tool, the Concordance, Validity, Safety and Accuracy (CVSA) model, is developed based on numerous established assessment tools.2,18 It provides an in-depth evaluation of answers generated by ChatGPT in terms of their concordance and homogeneity. Additionally, it scrutinizes the validity of the response in order to identify hallucinations, which are a major concern in the applications of LLMs in healthcare. Notably, it introduces a mechanism to report and flag responses that could potentially lead to unsafe or harmful practices for our patients.

This marks a significant stride towards addressing patient safety concerns in utilizing LLM in healthcare. To our knowledge, CVSA is the first of its kind designed to systematically evaluate LLM with a strong emphasis on patient safety.

The data was analyzed using Google Sheets and statistical analysis was performed using Microsoft Excel 2022.

## Results

The preliminary data with questions and responses can be found in Attachments 3 - 5.

For direct inquiries proposed to ChatGPT, the system achieved a cumulative score of 23.5 out of a possible 34, equaling 69.1%. The minimum passing score for Royal College Examination is 70%. Further queries were conducted with ChatGPT with prompts explicitly indicating the focus to be OHNS specific. Under these conditions, ChatGPT exhibited superior performance, achieving a score of 75.0% on the initial trial. When comparing the first attempt and the second attempt of ChatGPT, the first attempt performed slightly better than the second attempt. The accuracy rate was found to be 72.1% when the program was asked to regenerate its answers. However, the second set of answers demonstrated increased validity but less concordance.

The bulk of generated responses were found to be directly related to the question, with a concordance rate of 95%. Outliers in this instance were characterized by two divergent responses that were either self-contradictory or incongruous with the posed question. Overall, the majority (67%) of responses were deemed valid, corroborated by either broadly accepted facts, OHNS consensus, evidence-based data, scientific validation, or alignment with the opinions of OHNS senior staff. A subset of the responses (23.8%) contained partially invalid answers, with a minute fraction (3.2%) being deemed mostly invalid. It was observed that these statements lacked scientific validity, adherence to evidence-based principles, or acceptance by the OHNS society, known as hallucinations. There are some answers (3.2%) that are verbose but do not contain information that can be assessed objectively.

To evaluate if there is any significant difference amongst different groups, ANOVA analysis was performed using Microsoft Excel. It was found there were not significant differences amongst different groups. (F = 0.06, F crit =3.15, P-value = 0.93)

## Discussion

The data presented in this study represents the first assessment of LLM such as ChatGPT for OHNS specialty board examinations. It is also the first assessment of medical specialty board examinations with open-end questions. The questions are in alignment with Canada Royal College OHNS certifying exams. This methodology is congruent with that employed by the Canadian board exams and several other nations.

This study utilized an official sample exam, which was meticulously reviewed by educational leads within the specialty and provides a strong correlation with real examination materials and difficulty level. Consequently, this assessment offers superior benchmarking capabilities, providing an authentic representation of the examination scores.

The open-ended questions endeavour to mimic real-life clinical scenarios, where physicians are frequently confronted with open-ended questions, challenging their capacity to reason and draw conclusions. Most other evaluations of LLMs’ performance in ChatGPT are based on multiple-choice questions, showcasing the AI’s ability to identify and incorporate key topics and crucial information. However, this format falls short in assessing the breadth of knowledge and reasoning capabilities of the AI.

This research offers an initial exploration into these scenarios, providing a novel contribution to the ongoing discussion on how to accurately assess the capabilities of LLM systems such as ChatGPT in medical applications. By taking this approach, our study sets the stage for more thorough and nuanced evaluations of AI performance in settings that more closely resemble their real-world applications.

### 1. The concordance of answers generated by ChatGPT

Overall, ChatGPT demonstrated considerate concordance, or have explanations that affirm the answer or are directly related to the question. Conversely, a response is deemed as discordant when any segment of the explanation contradicts itself, or is not directly related to the question. This piece of our assessment tool is particularly useful for LLMs such as ChatGPT, which are known to generate large amounts of text data with low information density.

During the evaluation, it was observed that the answers provided by ChatGPT were generally concordant (92.06%) and directly addressed the questions posed. Only 9% of the response contained conflicting or verbose unrelated information. For instance, in one answer, ChatGPT incorrectly stated that the symptoms were solely caused by a bacterial infection, providing a lengthy explanation. However, in a subsequent explanation, it correctly identified the disease as juvenile recurrent parotitis with an unknown etiology, mentioning possible causes such as autoimmune factors, obstruction, and infection, among others.

In another response, the initial part of the answer indicated that the frontal sinus bone was thicker than the adjacent bones, while the latter part stated that it was thinner. This conflicting information demonstrates the lack of inherent understanding of the text by ChatGPT even though they were generated by itself.

### 2. The validity of answers generated by ChatGPT

The majority of the answers provided by ChatGPT were found to be valid. 66.67% were identified as valid, 23.81% were identified as mostly valid, and 9.52% were found to be indeterminate, fair, or mostly invalid, as demonstrated in Figure 1.

**Figure 1.**
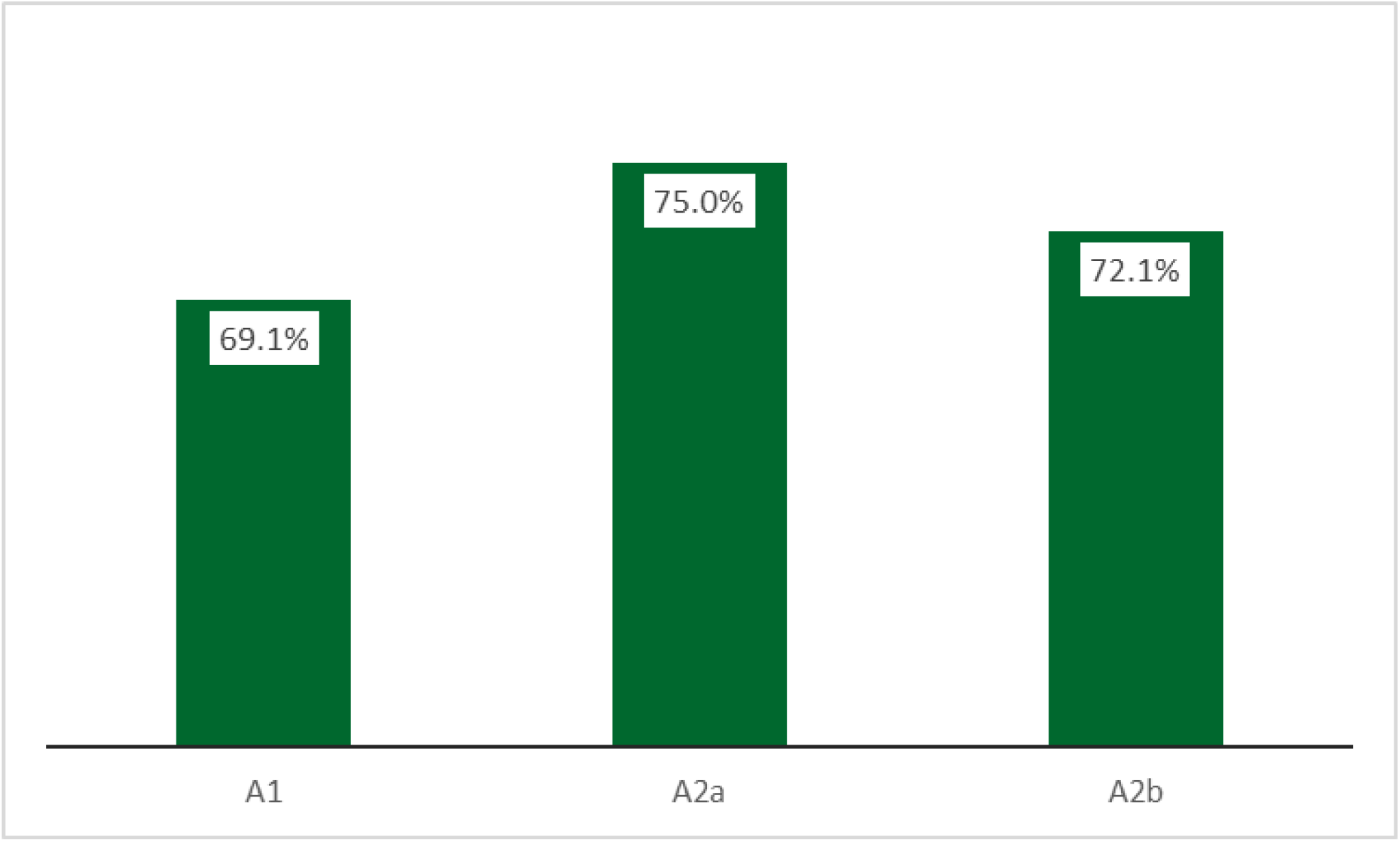
Accuracy comparison of 3 different groups. (A1: without prompt, A2: first attempt with prompt, A2b: second attempt with attempt)

**Figure 2.**
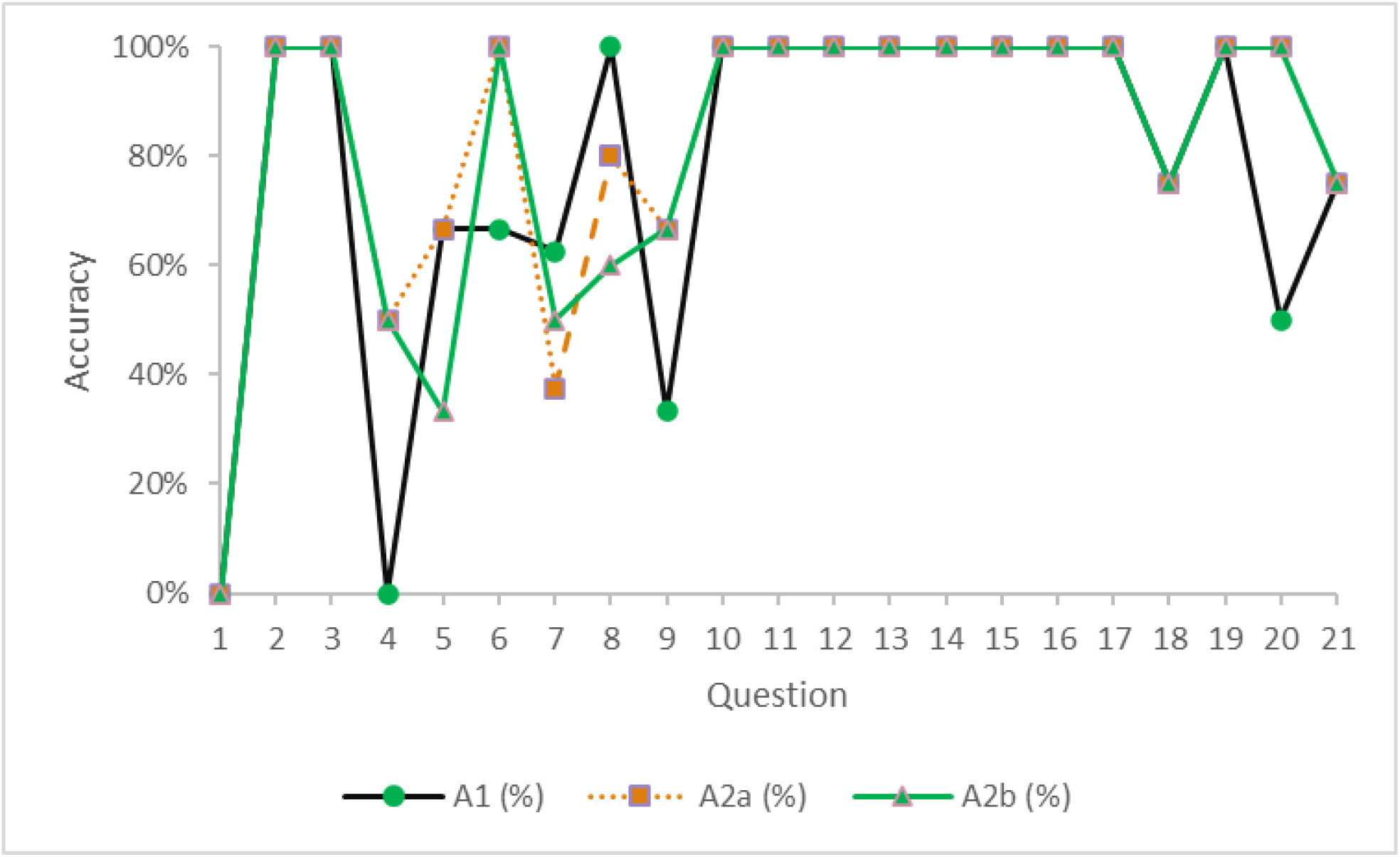
Scoring details of 3 different groups. (A1: without prompt, A2: first attempt with prompt, A2b: second attempt with attempt)

**Figure 3.**
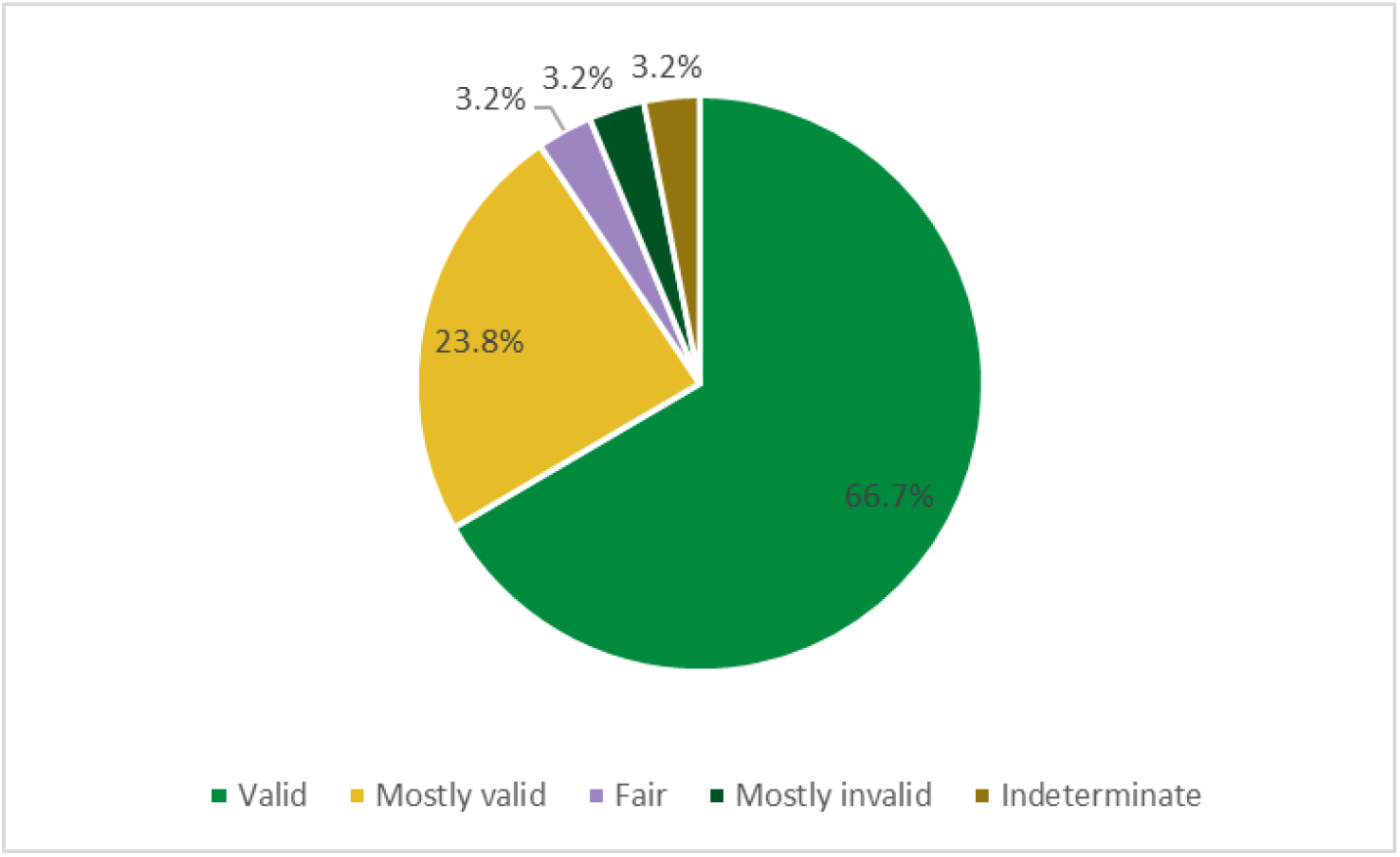
Overall validity of all answers combined.

**Figure 4.**
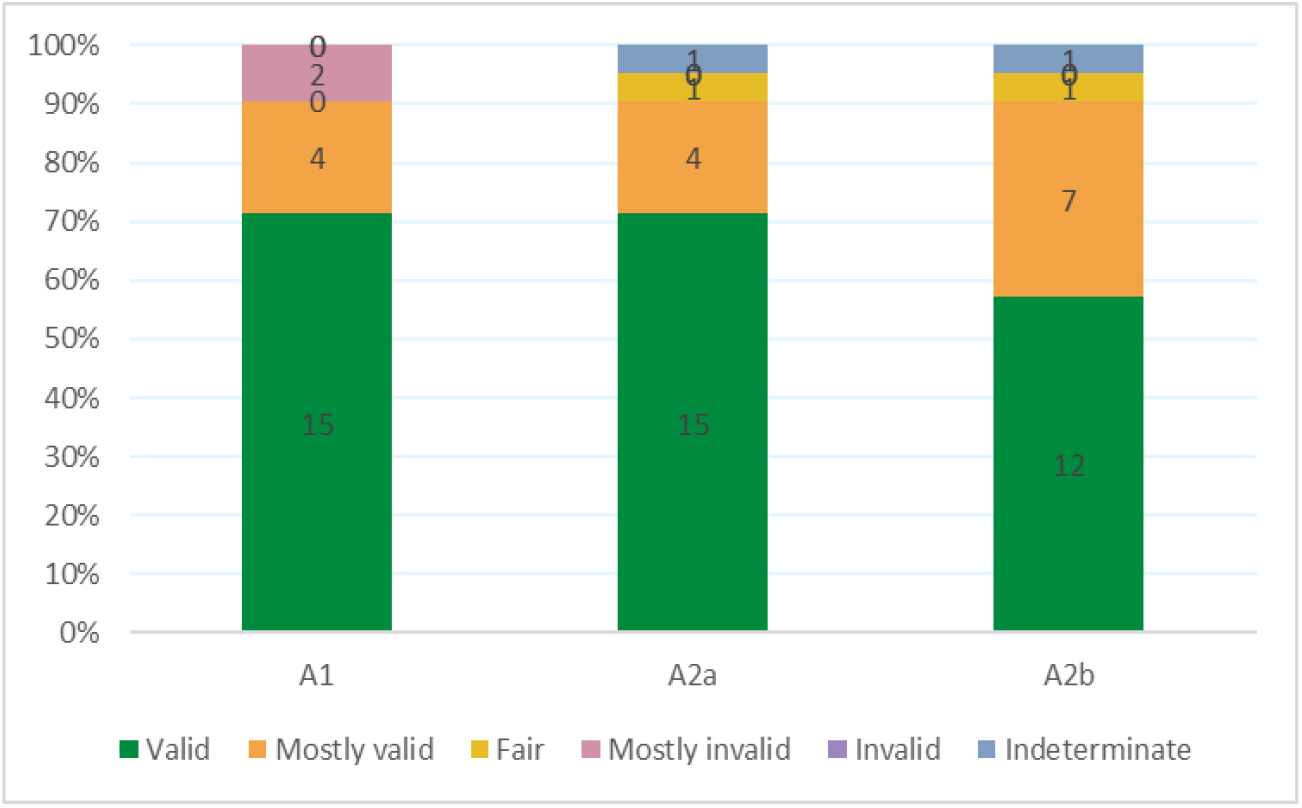
Validity of different groups. (A1: without prompt, A2: first attempt with prompt, A2b: second attempt with attempt)

Language Models (LMs), including ChatGPT, have been known to generate hallucinations, which are characterized by blatant factual errors, significant omissions, and erroneous information generation.^19^ The high linguistic fluency of LMs allows them to interweave inaccurate or unfounded opinions with accurate information, making it challenging to identify such hallucinations.

For example, in one of the answers, ChatGPT introduced the term “recurrent bacterial parotitis,” which is not a recognized diagnosis accepted by the OHNS society. Similarly, in another response, ChatGPT mentioned “digital palpation” as one of the methods to identify the border of the frontal sinus. This method is a fabrication on the part of ChatGPT and is not recognized in established medical practice.

Overall, it was observed that ChatGPT demonstrated a strong understanding of foundational anatomy and the pathophysiology of OHNS disease presentations. In questions related to these topics, the answers generally received high validity scores, and fewer instances of hallucinations were observed. It is possible that the extensive text data available on these subjects allowed the LLM to draw information and generate more accurate responses.

### 3. Patient safety concerns in the answers

Hallucinations may present benign or harmful information, with particular implications in the field of medicine. Such hallucinations could include misleading or incorrect data, and if followed by clinical practitioners, may pose substantial risks to patient safety. In our evaluation, we asked evaluators to identify and red flag such statements they encountered.

Certain hallucinations, albeit inaccurate, do not impact patient safety significantly. For instance, ChatGPT occasionally utilizes very outdated terminology. An example of this is the usage of “recurrent parotitis” rather than the full terms “juvenile recurrent parotitis” or “recurrent parotitis of childhood” which are currently widely accepted.

There are situations where ChatGPT’s inaccuracies can potentially compromise patient safety. For instance, when asked about the planes of a bicoronal approach for an osteoplastic flap, ChatGPT provided incorrect information, which could, in certain cases, jeopardize the health of the flap. Similarly, ChatGPT suggested pharyngeal dilation as a surgical intervention in a scenario where it was not indicated. This could place a patient at risk of undergoing unnecessary surgical procedures if the recommendation is followed precisely. Another instance of potentially harmful misinformation includes ChatGPT’s suggestion of laryngotracheal reconstruction for an anterior glottic web, an approach that is excessively radical for the condition.

### 4. The overall accuracy of the result

In our study, ChatGPT performed well and secured passing scores in all three tests: the unprompted test, the first attempt with a prompt, and the regenerated answer with a prompt, scoring 69.1%, 75.0%, and 72.1%, respectively.

Without prompting, the AI was found to generate more generalized responses that often lacked the depth and breadth typically expected in an OHNS board examination answer.

ChatGPT-4 demonstrated potential in successfully navigating complex surgical specialty board exams, specifically when presented with open-ended questions. Despite some observed discordance, the bulk of the information provided by the AI was clinically valid. Such features may prove highly beneficial for medical education, especially in low-income settings where access to such information may not be as readily available.

Some inaccuracies identified were due to the use of outdated data. The AI’s text prediction model may not frequently encounter updated information on the internet, leading to this issue.

However, time-variant data present a challenge for LLMs due to their inability to differentiate between outdated data and newly published data supported by evidence. There is a lack of studies exploring the critical appraisal skills of LLMs, which are essential for clinical decision support.

Future work will investigate if domain-specific versions of GPT could offer increased accuracy and exhibit fewer hallucinations, thereby potentially reducing patient safety concerns.

## Limitations

While this study presents valuable insights of the performance of ChatGPT in open end OHNS questions, it is important to acknowledge its inherent limitations:

1. Image-based questions could not be utilized for assessment due to the limitations of the currently available ChatGPT-4, which the public version does not support visual data queries at the time of our test. Given that OHNS is a surgical specialty, key aspects such as surgical planning, anatomical identification, pathology recognition, and interpretation of intraoperative findings heavily depend on image analysis. Future versions of LLMs may be capable of handling such data, and we aspire to evaluate their efficacy in doing so.
2. The study’s data collection and validation methods require a more extensive set of questions. Only 21 questions were adopted from the Royal College’s sample set for this study. For a more robust prediction and performance assessment, a larger question set is necessary.

## Conclusion

We evaluated the performance of ChatGPT-4 by employing it on a sample board-certifying exam of the Royal College of Physicians and Surgeons of Canada for OHNS, utilizing our novel CVSA framework. ChatGPT-4 achieved a passing score on the test, indicating its potential competence in this specialized field. Nevertheless, certain reservations persist, notably the potential risk to patient safety due to hallucinations. Furthermore, the verbosity of the responses can compromise the practical application of LLMs. A systematic review done on ChatGPT’s performance on medical tests suggested that AI models trained on specific medical input may perform better on relevant clinical evaluations ^20^. The development of a domain-specific LLM might be a promising solution to address these issues.

## Data Availability

All data produced in the present work are contained in the manuscript

## Acknowledgements

We thank Neil Saduka (Klavier AI) and Deepak Subburam (Klavier AI) for their assistance and contributions during the course of this research.

## Notes

### Competing Interest Statement

The authors have declared no competing interest.

### Funding Statement

This study did not receive any funding

## References

1. Varanasi, L. AI models like ChatGPT and GPT-4 are acing everything from the bar exam to AP Biology. Here’s a list of difficult exams both AI versions have passed. Business Insider (2023).

2. Kung, T. H. et al. Performance of ChatGPT on USMLE: Potential for AI-assisted medical education using large language models. PLOS Digit Health 2, e0000198 (2023).

3. Bommineni, V. L., Bhagwagar, S., Balcarcel, D., Davazitkos, C. & Boyer, D. Performance of ChatGPT on the MCAT: The road to personalized and equitable premedical learning. medRxiv (2023) doi :10.1101/2023.03.05.23286533.

4. Sallam, M. The utility of ChatGPT as an example of large language models in healthcare education, research and practice: Systematic review on the future perspectives and potential limitations. medRxiv 2023–2002 (2023).

5. Rudolph, J., Tan, S. & Tan, S. ChatGPT: Bullshit spewer or the end of traditional assessments in higher education? JALT 6, (2023).

6. Chen, S. et al. The utility of ChatGPT for cancer treatment information. medRxiv 2023.03.16.23287316 (2023) doi: 10.1101/2023.03.16.23287316.

7. Antaki, F., Touma, S., Milad, D., El-Khoury, J. & Duval, R. Evaluating the Performance of ChatGPT in Ophthalmology: An Analysis of its Successes and Shortcomings. Ophthalmology Science 100324 (2023).

8. Sinha, R. K., Deb Roy, A., Kumar, N. & Mondal, H. Applicability of ChatGPT in Assisting to Solve Higher Order Problems in Pathology. Cureus 15, e35237 (2023).

9. Ali, R. et al. Performance of ChatGPT and GPT-4 on neurosurgery written board examinations. bioRxiv (2023) doi: 10.1101/2023.03.25.23287743.

10. Ali, R., Tang, O. Y., Connolly, I. D., Fridley, J. S. & Shin, J. H. Performance of ChatGPT, GPT-4, and Google Bard on a Neurosurgery Oral Boards Preparation Question Bank. medRxiv (2023).

11. Nógrádi, B. et al. ChatGPT M.D.: Is There Any Room for Generative AI in Neurology and Other Medical Areas? (2023) doi: 10.2139/ssrn.4372965.

12. Gilson, A., Safranek, C., Huang, T., Socrates, V. & Chi, L. How does ChatGPT perform on the medical licensing exams. The Implications of Large.

13. Skalidis, I. et al. ChatGPT takes on the European Exam in Core Cardiology: an artificial intelligence success story? Eur Heart J Digit Health ztad029 (2023).

14. Nastasi, A. J., Courtright, K. R., Halpern, S. D. & Weissman, G. E. Does ChatGPT provide appropriate and equitable medical advice?: A vignette-based, clinical evaluation across care contexts. medRxiv (2023) doi: 10.1101/2023.02.25.23286451.

15. Lee, P., Bubeck, S. & Petro, J. Benefits, limits, and risks of GPT-4 as an AI chatbot for medicine. N. Engl. J. Med. 388, 1233–1239 (2023).

16. Format of the Examination in Vascular Surgery. https://www.royalcollege.ca/rcsite/documents/ibd/otolaryngology_examformat_e#:~:text=The%20Royal%20College%20examination%20in,you%20must%20pass%20both%20components.

17. Hiesinger, W. et al. Almanac: Retrieval-Augmented Language Models for Clinical Medicine. Res Sq (2023) doi: 10.21203/rs.3.rs-2883198/v1.

18. GPT-4. https://openai.com/product/gpt-4.

19. Nori, H., King, N., McKinney, S. M., Carignan, D. & Horvitz, E. Capabilities of GPT-4 on Medical Challenge Problems. (2023).

20. Li, J., Dada, A., Kleesiek, J. & Egger, J. ChatGPT in Healthcare: A Taxonomy and Systematic Review. medRxiv 2023–2003 (2023).

