## Supplement 1 for "Evaluating ChatGPT-4 in Otolaryngology–Head and Neck Surgery Board Examination using the CVSA Model"

| **Scenario no.** | **Scenario and questions** | **Model answer** |
| --- | --- | --- |
| 1 | A 6 year old boy is referred to you by his family physician. He has had 3 episodes of painful swelling of the right parotid gland over the past 18 months. Each episode lasted 5-7 days and has been accompanied by a low grade fever. Two of the episodes were managed with amoxicillin. One of the episodes was not treated with antibiotics and spontaneously resolved. There was complete resolution of symptoms between episodes. The child is otherwise healthy.   1. What is the most likely diagnosis? 2. List 4 treatment measures that are most commonly employed in the management of this condition. 3. What would you tell the parents about the long term clinical course of this disease? | 1. 1 mark, max 1 mark    a. Recurrent parotitis of childhood (must have entire term)    b. Also accept JRP (juvenile recurrent parotitis). 2. 0.5 marks for each, max 2 marks    a. Massage    b. Hydration    c. Analgesics    d. Antibiotics    e. Sialogogues    f. Heat (only 0.25 mark for this) 3. 1 mark each, max 1 mark    a. More recurrences    b. Probable remission as child enters adolescence    c. Sialoendoscopy |
| 2 | You are performing an osteoplastic flap procedure using a bicoronal approach for drainage of mucoceles.   1. In which soft tissue planes will you elevate the bicoronal flap? 2. Once the frontal bone is exposed, list three techniques to identify margins of the frontal sinus prior to making the bony cuts for the osteoplastic flap. | 1. 1 mark each, 2 marks max    a. Subgaleal plane in the area between superior temporal lines    b. Superficial layer of deep temporal fascia overlying temporalis muscle    c. NO marks for subperiosteal plane 2. 1 mark each, 3 marks max    a. Plain x-ray used as template    b. Intraoperative image guidance/surgical navigation    c. Transillumination via frontal trephine |
| 3 | List the three most common complications associated with placement of tympanostomy tubes in children. | 1 mark each, 3 marks max   a. Otorrhea b. Persistent perforation c. Granulation tissue formation |
| 4 | Name 4 local oncologic contraindications to supra-cricoid laryngectomy. | 1 mark each, 4 marks max   a. Fixation of the arytenoid cartilage secondary to cricoarytenoid joint fixation b. Subglottic extension to the level of the cricoid or direct invasion of the cricoid    i. Paraglottic extension c. Invasion of the posterior commissure (interarytenoid area) d. Extension of the outer perichondrium of the thyroid cartilage or extralaryngeal spread |
| 5 | A patient presents with an anterior glottic web. Name 5 treatment options, used alone or in combination, for the management of this lesion. | 0.5 marks each, 2.5 marks max   a. Observation b. CO_2_ laser c. Cold dissection d. Mitomycin C e. Laryngofissure with keel insertion f. Steroid injection g. Endoscopic keel |
| 6 | Name 3 techniques used to prevent or cure pharyngoesophageal spasm after total laryngectomy. | 1 mark each, 3 marks max   a. Cricopharyngeal myotomy b. Pharyngeal plexus neurectomy c. Botox injection |
| 7 | What is the main arterial supply to each of the following flaps once they are elevated?   1. Pectoralis major flap 2. Latissimus dorsi flap 3. Lower island trapezius flap 4. Paramedian forehead flap 5. Deltopectoral flap 6. Fibular free flap 7. Iliac osteomyocutaneous free flap | 0.5 marks each, 3.5 marks max   1. Pectoral branch of thoracoacromial    a. Lateral thoracic (0.25 marks only) 2. Thoracodorsal 3. Transverse cervical    a. Dorsal scapular (0.25 marks only) 4. Supratrochlear 5. Perforator intercostal of the internal mammary 6. Peroneal 7. Deep circumflex iliac |
| 8 | A 49 year old patient with T3N2b squamous cell carcinoma of the hypopharynx is scheduled to receive combined chemotherapy and radiation. What is the most important potential long-term sequelae of this treatment that must be disclosed to the patient? | 1 mark, 1 mark max   a. Permanent dysphagia/need for tube feeding    i. Airway obstruction or xerostomia (0.5 mark only) |
| 9 | List four genera of fungi that cause mucormycosis. | 0.5 mark each, 2 marks max   a. Mucor b. Rhizomucor c. Rhizopus d. Absidia e. Apophysomyces |
| 10 | 1. List 2 functions of the superior laryngeal nerve. 2. Where do the recurrent laryngeal nerve and the internal branch of the superior laryngeal nerve enter the larynx respectively? | 1. 0.5 marks each, 1 mark max    a. Internal branch - sensory to the mucosa of the larynx superior to the true vocal cord       i. *need to be specific as to where it provides sensation    b. External branch - motor to the cricothyroid muscle 2. 0.5 marks each, 1 mark max    a. Recurrent nerve enters posterior to the cricothyroid joint    b. Internal branch of the superior laryngeal nerve pierces the thyrohyoid membrane along with the superior laryngeal artery       i. *detail about artery as mentioned above not required to receive point |
| 11 | List all of the neural and vascular structures that pass through the sphenopalatine foramen. | 1 mark each, 4 marks max   a. Sphenopalatine artery b. Sphenopalatine vein c. Nasopalatine nerve d. Posterosuperior lateral nasal nerve |
