## Supplement 2 for "Evaluating ChatGPT-4 in Otolaryngology–Head and Neck Surgery Board Examination using the CVSA Model"

| Adjudication criteria: C-VS-A Scoring System | |
| --- | --- |
| Concordance  (Does AI understand the question, and can form a cohesive answer?) | - **Concordant**: Explanation affirms the answer or is directly related to the question. - **Discordant**: Any part of the explanation contradicts itself, or is not directly related to the question |
| Validity  (Of all the information presented by AI, how much of it are valid (widely-accepted opinion, OHNS consensus, evidence based, scientifically proven, or OHNS senior staff opinion)? ) | - **Valid**: All information given is valid - **Mostly valid**: ≥ 1 piece of information is invalid, but more than 50% of information presented is valid - **Fair**: Proportion of valid to invalid information is equal - **Mostly invalid**: ≥ 1 piece of information is valid, but more than 50% of information presented is invalid - **Invalid**: All information is invalid - **Indeterminate**: Contains generic information not applicable to given clinical scenario/question or does not answer the question |
|  | If chose any option other than valid in the above question, answer the follow question: |
|  | Does the answer contain information which is misleading, faulty or non-proven, that following the guidance of so, might compromise patient **safety**?   - Yes - provide detail in the comment box below - No |
| Accuracy  (The overall performance of AI, does it miss any important parts of the answer?) | Numeric score out of (*), points assigned according to answer key. |
