## Supplement 3 for "Evaluating ChatGPT-4 in Otolaryngology–Head and Neck Surgery Board Examination using the CVSA Model"

| **Scenario no.** | **Scenario and questions** | **Model answer** | **GPT-4 answers (without lead in and in a new session from A1 and A2)** |
| --- | --- | --- | --- |
| 1 | A 6 year old boy is referred to you by his family physician. He has had 3 episodes of painful swelling of the right parotid gland over the past 18 months. Each episode lasted 5-7 days and has been accompanied by a low grade fever. Two of the episodes were managed with amoxicillin. One of the episodes was not treated with antibiotics and spontaneously resolved. There was complete resolution of symptoms between episodes. The child is otherwise healthy.   1. What is the most likely diagnosis? 2. List 4 treatment measures that are most commonly employed in the management of this condition. 3. What would you tell the parents about the long term clinical course of this disease? | 1. 1 mark, max 1 mark    a. Recurrent parotitis of childhood (must have entire term)    b. Also accept JRP (juvenile recurrent parotitis). 2. 0.5 marks for each, max 2 marks    a. Massage    b. Hydration    c. Analgesics    d. Antibiotics    e. Sialogogues    f. Heat (only 0.25 mark for this) 3. 1 mark each, max 1 mark    a. More recurrences    b. Probable remission as child enters adolescence    c. Sialoendoscopy | **Q1, A3**: I am not a doctor, but based on the information provided, the most likely diagnosis for the 6-year-old boy with recurrent episodes of painful swelling of the right parotid gland, low grade fever, and resolution of symptoms between episodes could be recurrent parotitis. Recurrent parotitis is a condition characterized by inflammation and infection of the parotid gland, which is one of the major salivary glands. This condition is relatively rare in children, but it can occur.  However, it is important for the child to be evaluated by a medical professional, as there could be other potential causes of the symptoms, such as a salivary gland stone (sialolithiasis), an autoimmune condition, or other underlying issues. A thorough evaluation, including a physical examination and imaging studies, may be necessary to determine the exact cause of the symptoms and establish the appropriate treatment plan.  **Q2, A3**: I am not a doctor, but I can provide some general information on the treatment measures that are commonly employed in the management of recurrent parotitis in children. It is important to consult a healthcare professional for accurate diagnosis and tailored treatment recommendations. Common treatment measures for recurrent parotitis may include:  Antibiotics: As the condition can be caused by bacterial infection, oral antibiotics such as amoxicillin may be prescribed to manage the acute episodes. Antibiotic treatment should be determined by a healthcare professional based on the specific needs of the patient.  Analgesics and anti-inflammatory medications: Over-the-counter pain relievers (e.g., acetaminophen, ibuprofen) may be used to manage pain and inflammation during acute episodes. These medications should be used as directed by a healthcare professional.  Hydration and sialogogues: Encouraging fluid intake, and using substances that stimulate saliva production (sialogogues), such as sugar-free sour candies or lemon juice, can help promote salivary flow and reduce inflammation in the parotid gland.  Warm compresses and massage: Applying warm compresses to the affected area and gently massaging the parotid gland can help alleviate pain and promote drainage of the gland.  In some cases, if conservative treatments are not effective, more invasive interventions such as sialendoscopy (a minimally invasive procedure that involves inserting a small camera into the salivary duct to visualize and remove obstructions) or surgery may be considered. The appropriate treatment plan should be determined by a healthcare professional based on the individual needs and circumstances of the patient.  **Q3, A3**: I am not a doctor, but I can provide you with some general information about the long-term clinical course of recurrent parotitis in children. It is essential to consult a healthcare professional for accurate information and tailored advice.  Recurrent parotitis is a condition that generally has a variable clinical course. Some children may experience spontaneous resolution of the condition over time, while others may continue to have recurrent episodes. The frequency and severity of episodes can also vary between individuals.  In many cases, the recurrent episodes of parotitis tend to decrease in frequency and severity as the child grows older, and some children may eventually stop having episodes altogether. However, it is important to note that the course of the disease can be different for each child, and it is difficult to predict with certainty how the condition will progress.  Parents should be encouraged to monitor their child's symptoms closely, maintain good oral hygiene, and seek appropriate medical care during acute episodes. Regular follow-up with a healthcare professional is important to ensure appropriate management of the condition and to address any complications or concerns that may arise.  It is important for parents to discuss the specific situation with their child's healthcare professional, who can provide more accurate information on the long-term clinical course and management of the disease based on their child's individual circumstances. |
| 2 | You are performing an osteoplastic flap procedure using a bicoronal approach for drainage of mucoceles.   1. In which soft tissue planes will you elevate the bicoronal flap? 2. Once the frontal bone is exposed, list three techniques to identify margins of the frontal sinus prior to making the bony cuts for the osteoplastic flap. | 1. 1 mark each, 2 marks max    a. Subgaleal plane in the area between superior temporal lines    b. Superficial layer of deep temporal fascia overlying temporalis muscle    c. NO marks for subperiosteal plane 2. 1 mark each, 3 marks max    a. Plain x-ray used as template    b. Intraoperative image guidance/surgical navigation    c. Transillumination via frontal trephine | **Q1, A3**: I am not a surgeon, but I can provide you with some general information about the osteoplastic flap procedure using a bicoronal approach for the drainage of mucoceles. Keep in mind that you should consult a qualified medical professional for accurate and personalized advice.  During an osteoplastic flap procedure using a bicoronal approach, the soft tissue planes that are typically involved in the elevation of the bicoronal flap include:   1. Skin and subcutaneous tissue: The initial incision is made in the skin, extending from one temporal region to the other, usually following the hairline to minimize visible scarring. The subcutaneous tissue is then dissected to expose the underlying galea aponeurotica. 2. Galea aponeurotica: This is the dense fibrous layer covering the calvaria. Dissection is done in the plane between the galea aponeurotica and the loose areolar tissue to preserve the integrity of the galea and minimize the risk of injury to the frontal branches of the facial nerve. 3. Loose areolar tissue: The dissection continues in this plane, which lies between the galea aponeurotica and the pericranium. The loose areolar tissue contains emissary veins, which should be carefully managed to avoid bleeding. 4. Pericranium: The dissection proceeds anteriorly, elevating the pericranium from the underlying calvaria. Care should be taken to preserve the pericranium as much as possible, as it will be used for closure and to promote healing.   Once the soft tissue planes have been elevated, the surgeon can proceed with the osteoplastic flap technique to access the mucocele and perform the necessary drainage.  Remember, it is crucial to consult a qualified medical professional for specific guidance on surgical procedures.  **Q2, A3**: I am not a surgeon, but I can provide you with general information on techniques that may be used to identify the margins of the frontal sinus before making bony cuts for the osteoplastic flap. You should consult a qualified medical professional for accurate and personalized advice.  Once the frontal bone is exposed, there are several techniques that can be used to identify the margins of the frontal sinus:   1. Anatomical landmarks: Surgeons can use anatomical landmarks to approximate the location of the frontal sinus. These landmarks include the supraorbital rim, the glabella, and the frontozygomatic suture lines. The frontal sinus typically lies between these landmarks, but variations in size and shape can exist. 2. Preoperative imaging: Preoperative imaging, such as computed tomography (CT) scans, can provide a detailed view of the patient's frontal sinus anatomy. This information can be used during the surgery to accurately identify the margins of the frontal sinus. Some surgeons may also use intraoperative navigation systems to correlate the surgical field with the preoperative imaging data, ensuring precise localization of the sinus margins. 3. Intraoperative transillumination: This technique involves shining a light through the frontal bone and observing the transmission of light through the sinus. The light will appear brighter where the bone is thinner, allowing the surgeon to identify the boundaries of the frontal sinus. However, this method may be less reliable in cases where the sinus is filled with mucosal thickening or other pathologies.   Each of these techniques has its advantages and limitations. Surgeons may choose to use a combination of these methods to ensure accurate identification of the frontal sinus margins during the osteoplastic flap procedure. Remember, it is crucial to consult a qualified medical professional for specific guidance on surgical procedures. |
| 3 | List the three most common complications associated with placement of tympanostomy tubes in children. | 1 mark each, 3 marks max   a. Otorrhea b. Persistent perforation c. Granulation tissue formation | **A3**: Tympanostomy tubes, also known as ear tubes or ventilation tubes, are small tubes placed in the eardrum to help drain fluid and relieve pressure in the middle ear. While the procedure is generally safe and effective, there can be some complications. The three most common complications associated with the placement of tympanostomy tubes in children are:   1. Otorrhea: This refers to drainage or discharge from the ear, which can occur when there is an infection or inflammation in the middle ear. It is the most common complication and can be treated with antibiotic ear drops or oral antibiotics. 2. Tube obstruction or blockage: This can happen when the tube becomes blocked by debris, such as dried mucus or earwax. This may cause a decrease in the tube's effectiveness and may require a healthcare provider to clean or remove the obstruction. 3. Persistent tympanic membrane perforation: After the tube is removed or falls out, the eardrum usually heals on its own. However, in some cases, the eardrum may not close completely, leaving a persistent perforation. This can lead to hearing loss, recurrent infections, or the need for additional surgical intervention to repair the eardrum. |
| 4 | Name 4 local oncologic contraindications to supra-cricoid laryngectomy. | 1 mark each, 4 marks max   a. Fixation of the arytenoid cartilage secondary to cricoarytenoid joint fixation b. Subglottic extension to the level of the cricoid or direct invasion of the cricoid    i. Paraglottic extension c. Invasion of the posterior commissure (interarytenoid area) d. Extension of the outer perichondrium of the thyroid cartilage or extralaryngeal spread | **A3**: Supra-cricoid laryngectomy (SCL) is a partial laryngectomy procedure that aims to preserve laryngeal function and maintain the airway while treating laryngeal cancer. However, there are certain local oncologic contraindications that may preclude a patient from undergoing this procedure. Four such contraindications include:   1. Involvement of the cricoid cartilage: If the tumor has invaded the cricoid cartilage or extended to its full thickness, a supra-cricoid laryngectomy may not be feasible, as this structure is critical for maintaining the airway and swallowing function postoperatively. 2. Extensive subglottic extension: If the tumor has extended significantly below the glottis, typically more than 1 cm, it may be challenging to achieve complete resection and clear margins with SCL, necessitating a more extensive surgical procedure. 3. Involvement of both arytenoid cartilages: The arytenoid cartilages play a vital role in voice and swallowing function. If both arytenoids are involved by the tumor, supra-cricoid laryngectomy may not be the best option, as the patient may experience poor functional outcomes postoperatively. 4. Presence of extralaryngeal tumor spread: If the tumor has spread outside the larynx, particularly to adjacent structures such as the base of the tongue, the trachea, or the esophagus, supra-cricoid laryngectomy may not provide adequate tumor clearance, and a more extensive surgical approach may be necessary. |
| 5 | A patient presents with an anterior glottic web. Name 5 treatment options, used alone or in combination, for the management of this lesion. | 0.5 marks each, 2.5 marks max   a. Observation b. CO_2_ laser c. Cold dissection d. Mitomycin C e. Laryngofissure with keel insertion f. Steroid injection g. Endoscopic keel | **A3**: An anterior glottic web is a fibrous band of tissue that forms between the vocal folds, typically at the anterior commissure, which can lead to voice problems or airway obstruction. Treatment options for this condition can vary depending on the severity of the web and the patient's individual needs. Here are five treatment options that can be used alone or in combination for the management of an anterior glottic web:   1. Observation and voice therapy: In mild cases, conservative management with observation and voice therapy may be appropriate. Voice therapy can help patients improve their vocal function and compensate for the presence of the glottic web. 2. Endoscopic lysis: This minimally invasive procedure involves using a laryngoscope to visualize the web and cutting or laser vaporization to remove the tissue. This technique can be effective for thin webs, but recurrence is common if the underlying cause is not addressed. 3. Open surgical excision: In cases of thicker or more extensive glottic webs, open surgery may be necessary to remove the web and restore normal vocal fold function. This approach typically involves a laryngofissure, where the larynx is opened, and the web is excised with sutures placed to reconstruct the anterior commissure. 4. Keel placement: A silastic or metal keel can be placed between the vocal folds to prevent reformation of the web after its removal. The keel may be inserted endoscopically or during open surgery and is typically left in place for several weeks. 5. Adjuvant therapies: In some cases, additional treatments may be used to enhance the outcomes of the primary interventions. For example, corticosteroids may be administered to reduce inflammation and scarring, and mitomycin C, an antifibrotic agent, can be applied topically to the surgical site to reduce the risk of web recurrence.   The choice of treatment will depend on factors such as the severity of the web, the patient's overall health, and the surgeon's expertise. It is important to work closely with a laryngologist and a speech-language pathologist to determine the best course of action and ensure optimal outcomes. |
| 6 | Name 3 techniques used to prevent or cure pharyngoesophageal spasm after total laryngectomy. | 1 mark each, 3 marks max   a. Cricopharyngeal myotomy b. Pharyngeal plexus neurectomy c. Botox injection | **A3**:  ChatGPT  Pharyngoesophageal spasm is a potential complication that can occur after a total laryngectomy, a surgery that removes the larynx (voice box) due to cancer or other conditions. Although I'm not a medical expert, I can offer you some general information about techniques that might be used to prevent or manage pharyngoesophageal spasm after this surgery:   1. Swallowing therapy: A speech and swallowing therapist can work with the patient to develop and practice exercises that help improve muscle coordination and relaxation in the pharyngoesophageal area. This may involve learning new swallowing techniques, head and neck positioning, and strategies to minimize spasms. 2. Medications: Certain medications may be prescribed to help relax the muscles and reduce spasms. These may include antispasmodic medications (e.g., dicyclomine or hyoscyamine) or muscle relaxants (e.g., baclofen). These medications should be used under the guidance of a healthcare professional. 3. Botox injections: Botulinum toxin (Botox) injections can be administered by a qualified healthcare professional directly into the affected muscles of the pharyngoesophageal area. This can help to reduce muscle contractions and spasms, leading to improved swallowing function.   Remember that it is important to consult with a healthcare professional, such as a surgeon, oncologist, or speech therapist, for personalized advice and recommendations based on the specific needs of the patient. |
| 7 | What is the main arterial supply to each of the following flaps once they are elevated?   1. Pectoralis major flap 2. Latissimus dorsi flap 3. Lower island trapezius flap 4. Paramedian forehead flap 5. Deltopectoral flap 6. Fibular free flap 7. Iliac osteomyocutaneous free flap | 0.5 marks each, 3.5 marks max   1. Pectoral branch of thoracoacromial    a. Lateral thoracic (0.25 marks only) 2. Thoracodorsal 3. Transverse cervical    a. Dorsal scapular (0.25 marks only) 4. Supratrochlear 5. Perforator intercostal of the internal mammary 6. Peroneal 7. Deep circumflex iliac | **Q1, A3**: The main arterial supply to the pectoralis major flap, once elevated, is the thoracoacromial artery. The thoracoacromial artery is a branch of the axillary artery, and it supplies blood to the pectoralis major muscle as well as other surrounding structures. This artery is crucial to the viability of the pectoralis major flap when used for reconstructive surgery.  **Q2, A3**: The main arterial supply to the latissimus dorsi flap, once elevated, is the thoracodorsal artery. The thoracodorsal artery is a branch of the subscapular artery, which itself is a branch of the axillary artery. The thoracodorsal artery provides blood supply to the latissimus dorsi muscle and is essential for maintaining the viability of the latissimus dorsi flap when used in reconstructive surgery.  **Q3, A3**: The main arterial supply to the lower island trapezius flap, once elevated, is the transverse cervical artery, specifically its dorsal scapular branch. The transverse cervical artery originates from the thyrocervical trunk, which is a branch of the subclavian artery. The dorsal scapular branch of the transverse cervical artery supplies blood to the trapezius muscle and is crucial for the viability of the lower island trapezius flap when used in reconstructive surgery.  **Q4, A3**: The main arterial supply to the paramedian forehead flap, once elevated, is the supratrochlear artery. The supratrochlear artery is a branch of the ophthalmic artery, which itself originates from the internal carotid artery. The supratrochlear artery provides blood supply to the forehead skin and underlying musculature, making it essential for maintaining the viability of the paramedian forehead flap when used in reconstructive surgery.  **Q5, A3**: The main arterial supply to the deltopectoral flap, once elevated, is the internal mammary artery (also known as the internal thoracic artery). The internal mammary artery is a branch of the subclavian artery. It runs deep to the deltopectoral groove and provides perforating branches that supply blood to the overlying skin and subcutaneous tissue, which is crucial for maintaining the viability of the deltopectoral flap when used in reconstructive surgery. Additionally, the flap may also receive blood supply from the lateral thoracic artery and thoracoacromial artery.  **Q6, A3**: The main arterial supply to the fibular free flap, once elevated, is the peroneal artery. The peroneal artery is a branch of the tibial-fibular trunk, which originates from the popliteal artery. The peroneal artery provides blood supply to the fibular bone, surrounding muscles, and overlying skin, making it crucial for maintaining the viability of the fibular free flap when used in reconstructive surgery.  **Q7, A3**: The main arterial supply to the iliac osteomyocutaneous free flap, once elevated, is the deep circumflex iliac artery (DCIA). The deep circumflex iliac artery is a branch of the external iliac artery, which originates from the common iliac artery. The DCIA provides blood supply to the iliac bone, overlying skin, and the attached muscles, making it essential for maintaining the viability of the iliac osteomyocutaneous free flap when used in reconstructive surgery. |
| 8 | A 49 year old patient with T3N2b squamous cell carcinoma of the hypopharynx is scheduled to receive combined chemotherapy and radiation. What is the most important potential long-term sequelae of this treatment that must be disclosed to the patient? | 1 mark, 1 mark max   a. Permanent dysphagia/need for tube feeding    i. Airway obstruction or xerostomia (0.5 mark only) | **A3**: I am not a doctor, but I can try to provide some general information on this topic. For a patient with T3N2b squamous cell carcinoma of the hypopharynx receiving combined chemotherapy and radiation, there are several potential long-term side effects that may be important to discuss. One of the most important potential long-term sequelae of this treatment could be swallowing difficulties (dysphagia). This is due to the damage to the muscles and nerves responsible for swallowing that can occur from radiation therapy in the head and neck region.  Other potential long-term side effects that may be important to disclose to the patient include:  1. Xerostomia (dry mouth) due to damage to the salivary glands.  2. Dental problems, such as tooth decay and gum disease, which may be exacerbated by dry mouth.  3. Speech difficulties, which may result from damage to the muscles and nerves involved in speech.  4. Changes in taste and smell.  5. Trismus (difficulty opening the mouth) due to scarring or damage to the muscles and tissues of the jaw.  6. Fibrosis (scarring) of the soft tissues and skin in the treated area.  7. Late radiation toxicity, such as tissue necrosis or damage to the blood vessels in the treated area.  8. Increased risk of secondary cancers due to radiation exposure.  It is crucial for the patient to have a thorough discussion with their healthcare provider to understand the potential risks and benefits of the treatment, as well as possible side effects, and to make informed decisions about their care. |
| 9 | List four genera of fungi that cause mucormycosis. | 0.5 mark each, 2 marks max   a. Mucor b. Rhizomucor c. Rhizopus d. Absidia e. Apophysomyces | **A3**: Mucormycosis is a rare and severe fungal infection caused by fungi in the order Mucorales. Four genera of fungi that cause mucormycosis include:  1. Rhizopus: This is the most common cause of mucormycosis. Rhizopus oryzae and Rhizopus microsporus are two species associated with the infection.  2. Mucor: Mucor species are another significant cause of mucormycosis, with Mucor circinelloides being one of the most common species responsible for the infection.  3. Lichtheimia (formerly Absidia): Lichtheimia species, such as Lichtheimia corymbifera, can also lead to mucormycosis.  4. Cunninghamella: Cunninghamella bertholletiae is a species within this genus that has been implicated in causing mucormycosis, particularly in immunocompromised patients.  It is important to note that mucormycosis mainly affects immunocompromised individuals, such as those with uncontrolled diabetes, organ transplant recipients, or patients receiving chemotherapy. Prompt diagnosis and treatment are crucial, as the infection can progress rapidly and lead to severe complications. |
| 10 | 1. List 2 functions of the superior laryngeal nerve. 2. Where do the recurrent laryngeal nerve and the internal branch of the superior laryngeal nerve enter the larynx respectively? | 1. 0.5 marks each, 1 mark max    a. Internal branch - sensory to the mucosa of the larynx superior to the true vocal cord       i. *need to be specific as to where it provides sensation    b. External branch - motor to the cricothyroid muscle 2. 0.5 marks each, 1 mark max    a. Recurrent nerve enters posterior to the cricothyroid joint    b. Internal branch of the superior laryngeal nerve pierces the thyrohyoid membrane along with the superior laryngeal artery       i. *detail about artery as mentioned above not required to receive point | **Q1, A3**: The superior laryngeal nerve, a branch of the vagus nerve (cranial nerve X), has two main functions:  1. Sensory function: The internal branch of the superior laryngeal nerve provides sensory innervation to the laryngeal mucosa above the level of the vocal cords. This includes the epiglottis, aryepiglottic folds, and the arytenoids. It is responsible for transmitting sensations such as pain, temperature, and touch from the larynx to the brain.  2. Motor function: The external branch of the superior laryngeal nerve provides motor innervation to the cricothyroid muscle, which is responsible for stretching and tensing the vocal cords. This action changes the pitch of the voice by altering the tension of the vocal cords during phonation.  **Q2, A3**: The recurrent laryngeal nerve and the internal branch of the superior laryngeal nerve enter the larynx at different points:  1. Recurrent laryngeal nerve: This nerve enters the larynx by passing posteriorly around the inferior border of the cricoid cartilage, which is the lowermost cartilage of the larynx. It then ascends in the tracheoesophageal groove, supplying all the intrinsic muscles of the larynx except the cricothyroid muscle. In addition, the recurrent laryngeal nerve provides sensory innervation to the laryngeal mucosa below the level of the vocal cords.  2. Internal branch of the superior laryngeal nerve: The internal branch enters the larynx through the thyrohyoid membrane, which is a broad, fibrous sheet that connects the thyroid cartilage (largest laryngeal cartilage) to the hyoid bone (a U-shaped bone in the neck). The internal branch of the superior laryngeal nerve provides sensory innervation to the laryngeal mucosa above the level of the vocal cords. |
| 11 | List all of the neural and vascular structures that pass through the sphenopalatine foramen. | 1 mark each, 4 marks max   a. Sphenopalatine artery b. Sphenopalatine vein c. Nasopalatine nerve d. Posterosuperior lateral nasal nerve | **A3**: The sphenopalatine foramen is a small opening located in the posterior part of the nasal cavity where the sphenoid bone meets the palatine bone. It allows communication between the nasal cavity and the pterygopalatine fossa. The main neural and vascular structures passing through the sphenopalatine foramen include:  1. Nasopalatine nerve: This sensory nerve, also known as the long sphenopalatine nerve, is a branch of the pterygopalatine ganglion. It innervates the nasal septum and the anterior part of the hard palate.  2. Sphenopalatine artery: A branch of the maxillary artery, the sphenopalatine artery supplies blood to the nasal cavity, nasal septum, and adjacent structures. It has various branches, including the posterior lateral nasal branches and the posterior septal branches.  3. Sphenopalatine vein: This vein drains blood from the nasal cavity and the adjacent structures, and accompanies the sphenopalatine artery. It eventually drains into the pterygoid plexus or the facial vein.  These structures pass through the sphenopalatine foramen and contribute to the blood supply and innervation of the nasal cavity and surrounding areas. |
| ~~12~~ | ~~Theoretically, by what percentage will the Z-plasties shown in this referenced illustration increase scar length?~~  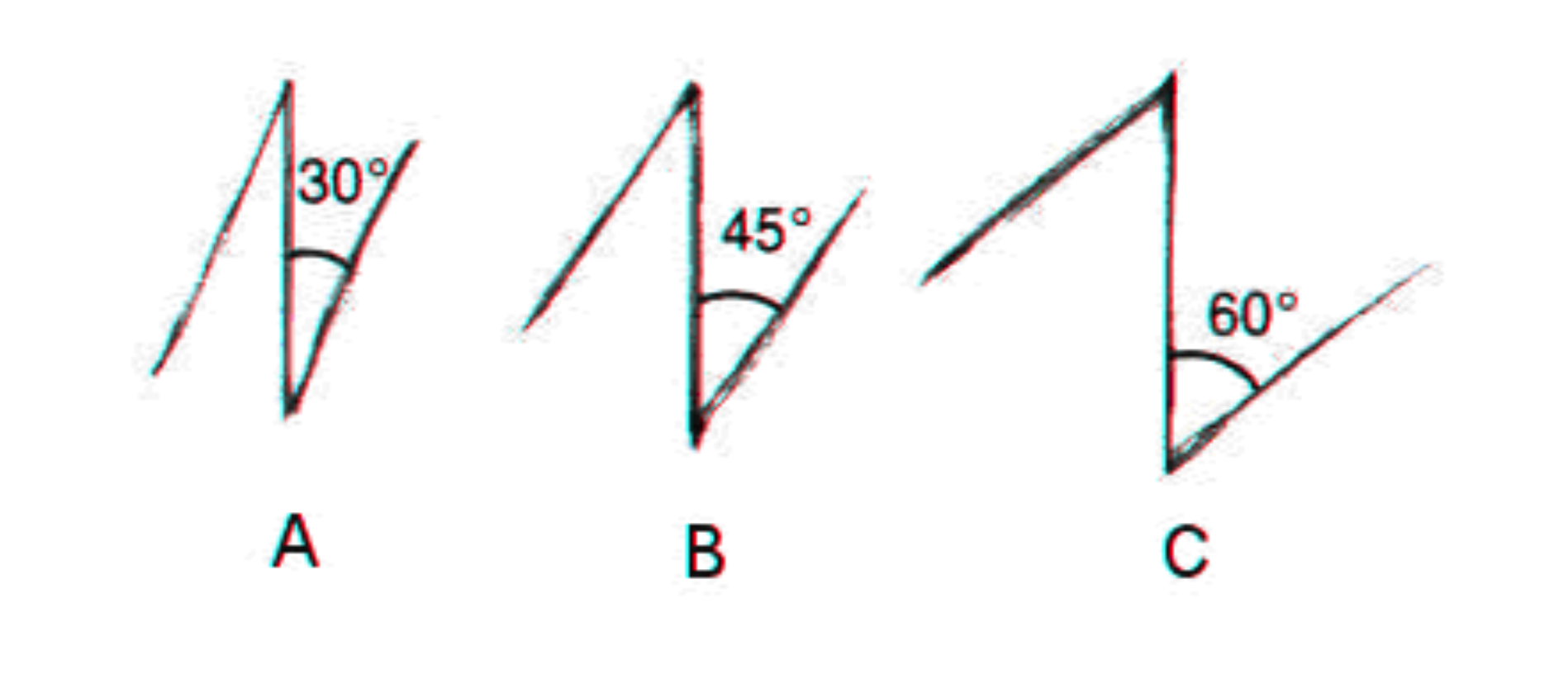   1. ~~A?~~ 2. ~~B?~~ 3. ~~C?~~ | 1. ~~25% (1 mark, max 1 mark)~~ 2. ~~50% (1 mark, max 1 mark)~~ 3. ~~75% (1 mark, max 1 mark)~~ |  |
| ~~13~~ | 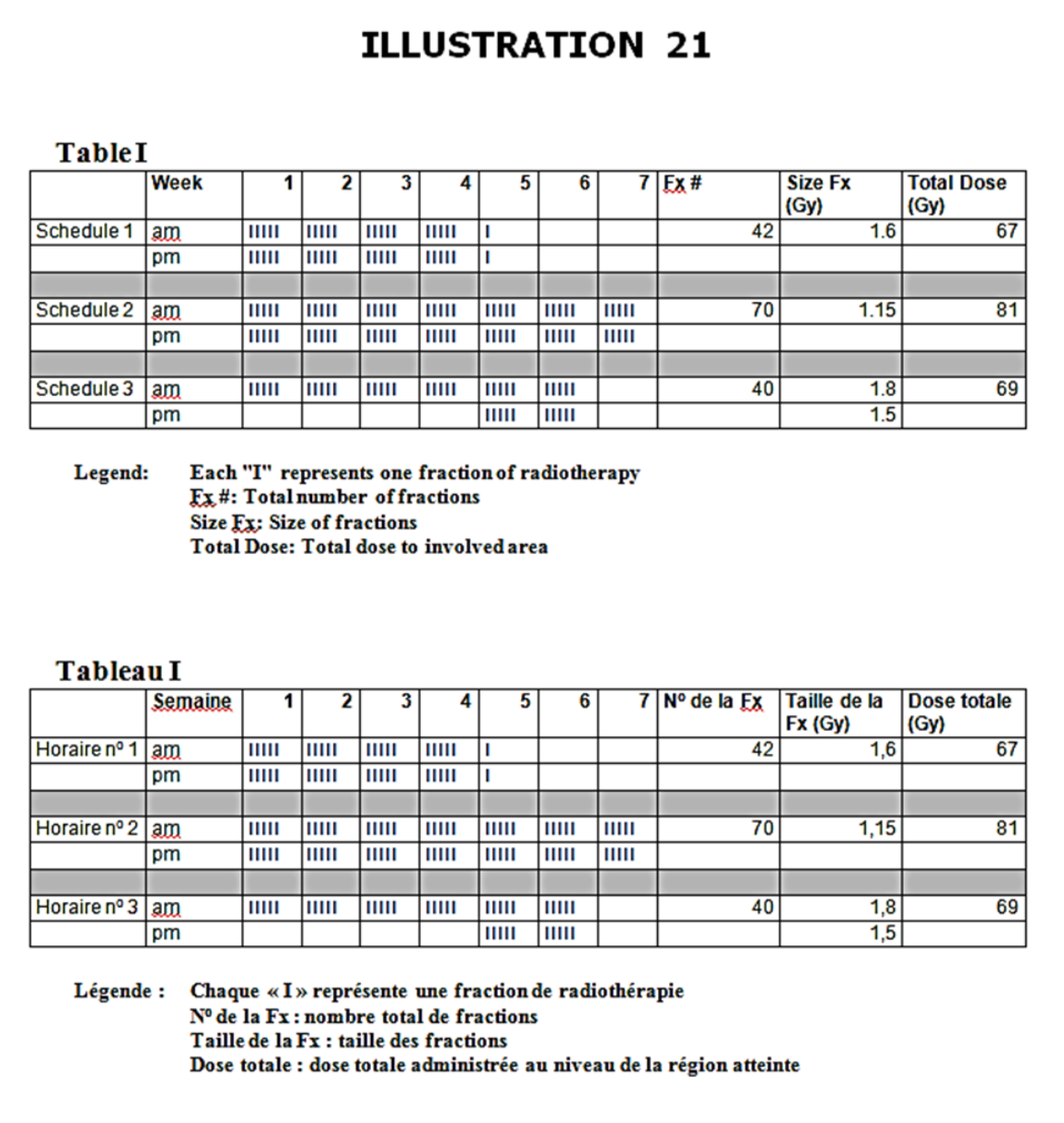  ~~See illustration.~~   1. ~~Name the types of radiation depicted in Table I.~~    a. ~~Schedule 1~~    b. ~~Schedule 2~~    c. ~~Schedule 3~~ 2. ~~What is the main radiobiologic rationale behind the type of radiation depicted in schedule 1?~~ 3. ~~What is the principal determinant of late radiation induced toxicity?~~ | ~~1 mark each, 5 marks max~~   1. a. ~~Accelerated~~    b. ~~Hyperfractionated~~    c. ~~Accelerated boost or concomitant boost~~ 2. ~~Reduce time for tumor to repopulate~~ 3. ~~Total dose~~ |  |
