## Supplement 4 for "Evaluating ChatGPT-4 in Otolaryngology–Head and Neck Surgery Board Examination using the CVSA Model"

| **Scenario no.** | **Scenario and questions** | **Model answer** | **GPT-4 answers (with lead in, used only in stem/prior to first question in set: “This is a question from an otolaryngology head and neck surgery licensing exam.”)** |
| --- | --- | --- | --- |
| 1 | A 6 year old boy is referred to you by his family physician. He has had 3 episodes of painful swelling of the right parotid gland over the past 18 months. Each episode lasted 5-7 days and has been accompanied by a low grade fever. Two of the episodes were managed with amoxicillin. One of the episodes was not treated with antibiotics and spontaneously resolved. There was complete resolution of symptoms between episodes. The child is otherwise healthy.   1. What is the most likely diagnosis? 2. List 4 treatment measures that are most commonly employed in the management of this condition. 3. What would you tell the parents about the long term clinical course of this disease? | 1. 1 mark, max 1 mark    a. Recurrent parotitis of childhood (must have entire term)    b. Also accept JRP (juvenile recurrent parotitis). 2. 0.5 marks for each, max 2 marks    a. Massage    b. Hydration    c. Analgesics    d. Antibiotics    e. Sialogogues    f. Heat (only 0.25 mark for this) 3. 1 mark each, max 1 mark    a. More recurrences    b. Probable remission as child enters adolescence    c. Sialoendoscopy | **Q1, A1**: The most likely diagnosis for this 6-year-old boy with recurrent episodes of painful swelling of the right parotid gland, accompanied by low-grade fever, is recurrent bacterial parotitis. Recurrent bacterial parotitis is an infection of the parotid gland that tends to occur in children, causing symptoms such as swelling, pain, and fever. It can be caused by various bacteria, with Staphylococcus aureus being one of the most common pathogens. The fact that the episodes were partially managed with amoxicillin suggests a bacterial etiology. Additionally, one episode resolved spontaneously, which can happen in some cases of bacterial parotitis.  It is important for the child to be evaluated by an otolaryngologist to confirm the diagnosis, determine the cause, and establish an appropriate treatment plan, which may include antibiotics and supportive care. Further investigation may be necessary to rule out other potential causes, such as sialolithiasis (salivary stones) or structural abnormalities that could predispose the child to recurrent infections.  **Q2, A1**: The management of recurrent parotitis in children typically involves a combination of conservative and medical treatments. Four commonly employed treatment measures include:   1. Warm compresses: Applying warm, moist heat to the affected area can help reduce pain and swelling by promoting blood flow and salivary gland secretion. 2. Hydration: Encouraging the child to drink plenty of fluids can help stimulate salivary flow, which may help flush out any potential blockage or infection. 3. Sialagogues: These are substances that help stimulate saliva production. Examples include sugar-free sour candies or lemon juice, which can be given to the child to encourage salivation and potentially help clear any blockages. 4. Antibiotics: If an infection is suspected or confirmed, antibiotics such as amoxicillin may be prescribed to treat the underlying bacterial infection. The choice of antibiotic may be guided by culture and sensitivity results if available.   It is important to note that these measures mainly address the symptoms and may not resolve the underlying cause. In cases where conservative and medical treatments are not effective, or if the cause is found to be an anatomical issue, further interventions such as sialography, ultrasound-guided needle aspiration, or surgical management (e.g., ductal dilation, ductoplasty, or parotidectomy) may be considered.  **Q3, A1**: When discussing the long-term clinical course of recurrent parotitis with the parents, it is important to provide a balanced and realistic outlook. You could tell them the following:   1. The course of the disease can vary from child to child. Some children may experience fewer and less severe episodes over time, while others may continue to have recurrent episodes. 2. Many children with recurrent parotitis eventually outgrow the condition as they get older, and the episodes may become less frequent or cease altogether. This is particularly true for cases of juvenile recurrent parotitis, which often resolves spontaneously during adolescence. 3. Maintaining good oral hygiene and staying hydrated can help reduce the risk of further episodes. Regular dental check-ups and following a healthy diet can also be beneficial for overall oral health. 4. In some cases, particularly if the cause is due to an anatomical issue or persistent obstruction, further evaluation and more definitive treatments, such as surgery, may be necessary. 5. It is important to closely monitor the child's condition and follow up with the healthcare provider for regular assessments and any necessary adjustments to the treatment plan. 6. Although recurrent parotitis can be uncomfortable and distressing for the child, it generally does not lead to severe long-term complications or significantly impact their overall health. However, it is crucial to continue to work with the healthcare provider to manage the condition and address any underlying issues. |
| 2 | You are performing an osteoplastic flap procedure using a bicoronal approach for drainage of mucoceles.   1. In which soft tissue planes will you elevate the bicoronal flap? 2. Once the frontal bone is exposed, list three techniques to identify margins of the frontal sinus prior to making the bony cuts for the osteoplastic flap. | 1. 1 mark each, 2 marks max    a. Subgaleal plane in the area between superior temporal lines    b. Superficial layer of deep temporal fascia overlying temporalis muscle    c. NO marks for subperiosteal plane 2. 1 mark each, 3 marks max    a. Plain x-ray used as template    b. Intraoperative image guidance/surgical navigation    c. Transillumination via frontal trephine | **Q1, A1**: In an osteoplastic flap procedure using a bicoronal approach for drainage of mucoceles, you will elevate the bicoronal flap in the following soft tissue planes:   1. Subgaleal plane: The incision is made in the scalp, and the dissection proceeds in the subgaleal plane, which is the space between the galea aponeurotica and the pericranium. This plane allows for the elevation of the scalp and galea together, preserving the blood supply and minimizing the risk of scalp necrosis. 2. Subperiosteal plane: Once the subgaleal dissection is completed to the level of the supraorbital rims and the lateral canthus, the dissection continues in the subperiosteal plane. This involves elevating the pericranium off the frontal bone and exposing the underlying bone. This plane is crucial for preserving the supraorbital and supratrochlear nerves, as well as the supraorbital vessels.   During the elevation of the bicoronal flap, care should be taken to preserve important neurovascular structures, such as the supraorbital and supratrochlear nerves and vessels.  **Q2, A1**: Once the frontal bone is exposed during an osteoplastic flap procedure, it is crucial to accurately identify the margins of the frontal sinus before making the bony cuts. Three techniques to help with this process are:   1. Digital palpation: One of the simplest methods to identify the margins of the frontal sinus is by using digital palpation. By carefully palpating the frontal bone, you can often feel the transition between the thicker bone of the frontal sinus and the thinner bone of the calvarial vault. The borders of the frontal sinus can be marked based on these tactile findings. 2. Preoperative imaging: Utilizing preoperative imaging, such as computed tomography (CT) scans, can provide a detailed anatomical roadmap of the patient's frontal sinus. By reviewing these images and noting the dimensions and borders of the frontal sinus, a surgeon can have a clearer understanding of the anatomy before making the bony cuts. Intraoperative image guidance systems can also be used to help with real-time visualization and localization of the frontal sinus margins. 3. Transillumination: This technique involves using a light source to shine through the thin bone overlying the frontal sinus. The light will appear brighter in areas where the bone is thinner, such as the frontal sinus, compared to the thicker bone of the calvarial vault. By transilluminating the frontal bone, you can visually identify the margins of the frontal sinus and mark them accordingly.   By using a combination of these techniques, a surgeon can accurately identify the margins of the frontal sinus and make precise bony cuts during the osteoplastic flap procedure. |
| 3 | List the three most common complications associated with placement of tympanostomy tubes in children. | 1 mark each, 3 marks max   a. Otorrhea b. Persistent perforation c. Granulation tissue formation | **A1**: The three most common complications associated with placement of tympanostomy tubes in children are:  Otorrhea: This refers to drainage or discharge from the ear, which can occur as a result of infection or a non-infectious process. Otorrhea is relatively common following tympanostomy tube placement, especially in the first few weeks after the procedure.  Tube obstruction or blockage: Obstruction of the tympanostomy tube can occur due to the accumulation of debris, such as cerumen (earwax), or a buildup of dried secretions. This may lead to reduced ventilation of the middle ear, which can cause hearing difficulties or recurrent ear infections.  Premature extrusion or dislodgment of the tube: Tympanostomy tubes can sometimes be dislodged or extrude prematurely from the eardrum. This can lead to inadequate ventilation of the middle ear or a need for additional procedures to reinsert the tube.  Other potential complications include persistent tympanic membrane perforation, granulation tissue formation, and hearing loss. However, these are less common than the three listed above. |
| 4 | Name 4 local oncologic contraindications to supra-cricoid laryngectomy. | 1 mark each, 4 marks max   a. Fixation of the arytenoid cartilage secondary to cricoarytenoid joint fixation b. Subglottic extension to the level of the cricoid or direct invasion of the cricoid    i. Paraglottic extension c. Invasion of the posterior commissure (interarytenoid area) d. Extension of the outer perichondrium of the thyroid cartilage or extralaryngeal spread | **A1**: Supra-cricoid laryngectomy is a surgical procedure used to treat laryngeal cancer while preserving the patient's voice and swallowing functions. However, there are local oncologic contraindications to this procedure, which make it unsuitable for certain cases. Four such contraindications include:   1. Involvement of the pre-epiglottic space: If the tumor has extended into the pre-epiglottic space, it increases the risk of recurrence and decreases the likelihood of a successful supra-cricoid laryngectomy. In such cases, a more extensive surgery might be necessary. 2. Involvement of the cricoid cartilage: Tumor invasion into the cricoid cartilage may compromise the structural integrity of the larynx, making it difficult or impossible to reconstruct after supra-cricoid laryngectomy. This contraindication may necessitate a total laryngectomy or alternative treatment options. 3. Involvement of the contralateral arytenoid: If the tumor has spread to the contralateral arytenoid cartilage (the arytenoid on the opposite side of the larynx), the patient may not be a suitable candidate for supra-cricoid laryngectomy, as the preservation of at least one arytenoid is crucial for the functional outcomes of the surgery. 4. Advanced T-stage tumors: Supra-cricoid laryngectomy is typically not recommended for patients with T3 or T4 tumors due to the extent of the disease and the increased risk of local recurrence. In these cases, a more aggressive surgical approach, such as total laryngectomy, or alternative treatments like chemoradiation might be considered.   It's important to note that each patient's situation is unique, and the presence of these contraindications should be considered in the context of the individual's overall health and treatment goals. |
| 5 | A patient presents with an anterior glottic web. Name 5 treatment options, used alone or in combination, for the management of this lesion. | 0.5 marks each, 2.5 marks max   a. Observation b. CO_2_ laser c. Cold dissection d. Mitomycin C e. Laryngofissure with keel insertion f. Steroid injection g. Endoscopic keel | **A1**: An anterior glottic web is a fibrous band of tissue that forms between the vocal folds and can lead to voice changes, breathing difficulties, and other issues. There are several treatment options that can be used alone or in combination for the management of this lesion. Five common options include:   1. Endoscopic lysis: This minimally invasive procedure involves using a laryngoscope to visualize the web and microsurgical instruments to carefully cut and separate the web. 2. Laser surgery: A CO2 or KTP laser can be used to vaporize the web and release the vocal folds. This approach can minimize bleeding and reduce the risk of scarring. 3. Laryngoplasty: A surgical procedure that involves reconstructing the larynx by removing the web and reshaping the affected area to restore normal vocal fold function. 4. Anterior commissure stenting: Following the removal of the web, a stent can be placed in the anterior commissure to prevent reformation of the web and promote healing of the vocal folds. 5. Voice therapy: In some cases, voice therapy may be used in conjunction with other treatments to help improve voice quality and function. This may involve exercises to improve vocal fold vibration, control pitch and loudness, and reduce strain on the larynx.   The choice of treatment depends on the severity of the web, the patient's symptoms, and the surgeon's experience and preference. In some cases, a combination of treatments may be necessary to achieve the best outcomes. |
| 6 | Name 3 techniques used to prevent or cure pharyngoesophageal spasm after total laryngectomy. | 1 mark each, 3 marks max   a. Cricopharyngeal myotomy b. Pharyngeal plexus neurectomy c. Botox injection | **A1**: Pharyngoesophageal spasm after total laryngectomy can lead to complications such as dysphagia and can affect the patient's quality of life. Several techniques can be used to prevent or cure pharyngoesophageal spasm after total laryngectomy. Here are three common techniques:   1. Myotomy: This involves the surgical division of the cricopharyngeal muscle to reduce the muscle's spasmodic action. This can help improve the passage of food or liquids through the pharynx and esophagus and relieve symptoms of pharyngoesophageal spasm. 2. Botulinum toxin injection: Botulinum toxin (Botox) can be injected into the cricopharyngeal muscle to temporarily weaken or paralyze it, reducing spasm and promoting smooth passage of food and liquids. This treatment is minimally invasive and can be repeated as necessary. 3. Pharyngeal dilation: This procedure involves the insertion of a dilator into the pharynx to gradually stretch the narrowed or spasmodic area, allowing for improved swallowing. This can be performed either through a rigid or a balloon dilator, depending on the patient's specific needs.   It is important to note that the choice of technique may depend on the severity of the spasm, the underlying cause, and the patient's overall health. Proper evaluation and consultation with an otolaryngologist is essential to determine the most appropriate treatment. |
| 7 | What is the main arterial supply to each of the following flaps once they are elevated?   1. Pectoralis major flap 2. Latissimus dorsi flap 3. Lower island trapezius flap 4. Paramedian forehead flap 5. Deltopectoral flap 6. Fibular free flap 7. Iliac osteomyocutaneous free flap | 0.5 marks each, 3.5 marks max   1. Pectoral branch of thoracoacromial    a. Lateral thoracic (0.25 marks only) 2. Thoracodorsal 3. Transverse cervical    a. Dorsal scapular (0.25 marks only) 4. Supratrochlear 5. Perforator intercostal of the internal mammary 6. Peroneal 7. Deep circumflex iliac | **Q1, A1**: The main arterial supply to a pectoralis major flap once it is elevated is the thoracoacromial artery, a branch of the axillary artery. This artery supplies blood to the pectoralis major muscle, making it an essential component of the flap's blood supply.  **Q2, A1**: The main arterial supply to the latissimus dorsi flap, once elevated, is the thoracodorsal artery. The thoracodorsal artery is a branch of the subscapular artery, which in turn arises from the axillary artery. The thoracodorsal artery provides blood supply to the latissimus dorsi muscle, which is the primary component of this flap.  **Q3, A1**: The main arterial supply to the lower island trapezius flap, once elevated, is the transverse cervical artery. The transverse cervical artery arises from the thyrocervical trunk, which is a branch of the subclavian artery. The transverse cervical artery provides blood supply to the trapezius muscle, which is the primary component of this flap. Specifically, the descending branch of the transverse cervical artery supplies the lower portion of the trapezius muscle used in the lower island trapezius flap.  **Q4, A1**: The main arterial supply to the paramedian forehead flap, once elevated, is the supratrochlear artery. The supratrochlear artery is a branch of the ophthalmic artery, which itself arises from the internal carotid artery. The supratrochlear artery provides blood supply to the forehead skin and underlying tissues, making it a reliable source of vascularization for the paramedian forehead flap.  **Q5, A1**: The main arterial supply to the deltopectoral flap, once elevated, is the internal mammary artery (also known as the internal thoracic artery). The internal mammary artery arises from the subclavian artery and runs deep to the clavicle, between the pectoralis major and deltoid muscles. Additionally, perforating branches from the lateral thoracic and thoracoacromial arteries can contribute to the vascularization of the flap.  **Q6, A1**: The main arterial supply to the fibular free flap, once elevated, is the peroneal artery, which is a branch of the tibial-fibular trunk (also known as the tibioperoneal trunk). The tibial-fibular trunk is a continuation of the popliteal artery in the lower leg. The peroneal artery provides blood supply to the fibula, the overlying skin, and the muscles in the lateral compartment of the leg, which are the primary components of the fibular free flap used in reconstructive surgeries.  **Q7, A1**: The main arterial supply to the iliac osteomyocutaneous free flap, once elevated, is the deep circumflex iliac artery (DCIA). The DCIA is a branch of the external iliac artery, which arises from the common iliac artery. The deep circumflex iliac artery provides blood supply to the iliac crest bone, as well as the overlying skin and the iliacus muscle, making it the primary vascular source for the iliac osteomyocutaneous free flap used in reconstructive procedures. |
| 8 | A 49 year old patient with T3N2b squamous cell carcinoma of the hypopharynx is scheduled to receive combined chemotherapy and radiation. What is the most important potential long-term sequelae of this treatment that must be disclosed to the patient? | 1 mark, 1 mark max   a. Permanent dysphagia/need for tube feeding    i. Airway obstruction or xerostomia (0.5 mark only) | **A1**: The most important potential long-term sequelae of combined chemotherapy and radiation for a patient with T3N2b squamous cell carcinoma of the hypopharynx are the development of radiation-induced dysphagia and aspiration. These complications can occur due to damage to the muscles and nerves involved in swallowing, leading to difficulty in swallowing solid foods and liquids, and an increased risk of inhaling food or liquid into the lungs. As a result, the patient may require a feeding tube or long-term nutritional support, and may be at risk of pneumonia or other respiratory complications. Other potential long-term sequelae of this treatment may include radiation-induced mucositis, xerostomia (dry mouth), hearing loss, and thyroid dysfunction, among others. However, the risk of dysphagia and aspiration is considered the most significant and therefore must be disclosed to the patient. |
| 9 | List four genera of fungi that cause mucormycosis. | 0.5 mark each, 2 marks max   a. Mucor b. Rhizomucor c. Rhizopus d. Absidia e. Apophysomyces | **A1**: Mucormycosis, also known as zygomycosis, is a rare and severe fungal infection caused primarily by fungi belonging to the order Mucorales. Four genera of fungi that are commonly associated with mucormycosis include:   1. Rhizopus 2. Mucor 3. Lichtheimia (previously known as Absidia) 4. Cunninghamella   These fungi are typically found in soil, decaying organic matter, and in the environment. They can cause a range of infections in humans, particularly in immunocompromised individuals, such as those with diabetes, organ transplant recipients, or patients receiving immunosuppressive therapy. Infections can manifest as rhino-orbital-cerebral, pulmonary, cutaneous, gastrointestinal, or disseminated mucormycosis, depending on the site of entry and spread of the fungi. |
| 10 | 1. List 2 functions of the superior laryngeal nerve. 2. Where do the recurrent laryngeal nerve and the internal branch of the superior laryngeal nerve enter the larynx respectively? | 1. 0.5 marks each, 1 mark max    a. Internal branch - sensory to the mucosa of the larynx superior to the true vocal cord       i. *need to be specific as to where it provides sensation    b. External branch - motor to the cricothyroid muscle 2. 0.5 marks each, 1 mark max    a. Recurrent nerve enters posterior to the cricothyroid joint    b. Internal branch of the superior laryngeal nerve pierces the thyrohyoid membrane along with the superior laryngeal artery       i. *detail about artery as mentioned above not required to receive point | **Q1, A1**: The superior laryngeal nerve, a branch of the vagus nerve (cranial nerve X), has two main functions:   1. Sensory: It provides sensory innervation to the laryngeal mucosa above the vocal cords. This is primarily done by its internal branch, the internal laryngeal nerve, which carries sensory information from the supraglottic region, including the epiglottis, aryepiglottic folds, and the upper part of the larynx. 2. Motor: It provides motor innervation to the cricothyroid muscle, which is responsible for lengthening and tensing the vocal cords, thereby increasing pitch. This function is performed by its external branch, the external laryngeal nerve, which supplies the cricothyroid muscle.   **Q2, A1**: The recurrent laryngeal nerve and the internal branch of the superior laryngeal nerve enter the larynx at different locations:   1. Recurrent laryngeal nerve: The recurrent laryngeal nerve enters the larynx by passing posterior to the cricothyroid joint and then underneath the inferior constrictor muscle. It provides motor innervation to all the intrinsic muscles of the larynx except the cricothyroid muscle, as well as sensory innervation to the laryngeal mucosa below the vocal cords. 2. Internal branch of the superior laryngeal nerve: The internal branch of the superior laryngeal nerve enters the larynx through the thyrohyoid membrane, which is located between the thyroid cartilage and the hyoid bone. It pierces the membrane superior to the superior thyroid cornu and provides sensory innervation to the laryngeal mucosa above the vocal cords. |
| 11 | List all of the neural and vascular structures that pass through the sphenopalatine foramen. | 1 mark each, 4 marks max   a. Sphenopalatine artery b. Sphenopalatine vein c. Nasopalatine nerve d. Posterosuperior lateral nasal nerve | **A1**: The sphenopalatine foramen is a small opening located in the posterior part of the superior nasal cavity. It connects the nasal cavity to the pterygopalatine fossa, allowing for the passage of both neural and vascular structures. The following structures pass through the sphenopalatine foramen:   1. Nasopalatine nerve (also known as the sphenopalatine nerve): a branch of the maxillary nerve (V2), which is the second division of the trigeminal nerve (cranial nerve V). 2. Sphenopalatine artery: a branch of the maxillary artery, which is one of the terminal branches of the external carotid artery. The sphenopalatine artery supplies blood to the nasal cavity and some parts of the oral cavity. 3. Sphenopalatine vein: a tributary of the pterygoid plexus that drains blood from the nasal cavity. 4. These structures are essential for the sensory and blood supply to the nasal cavity and play a vital role in the function of the upper respiratory tract. |
| ~~12~~ | ~~Theoretically, by what percentage will the Z-plasties shown in this referenced illustration increase scar length?~~  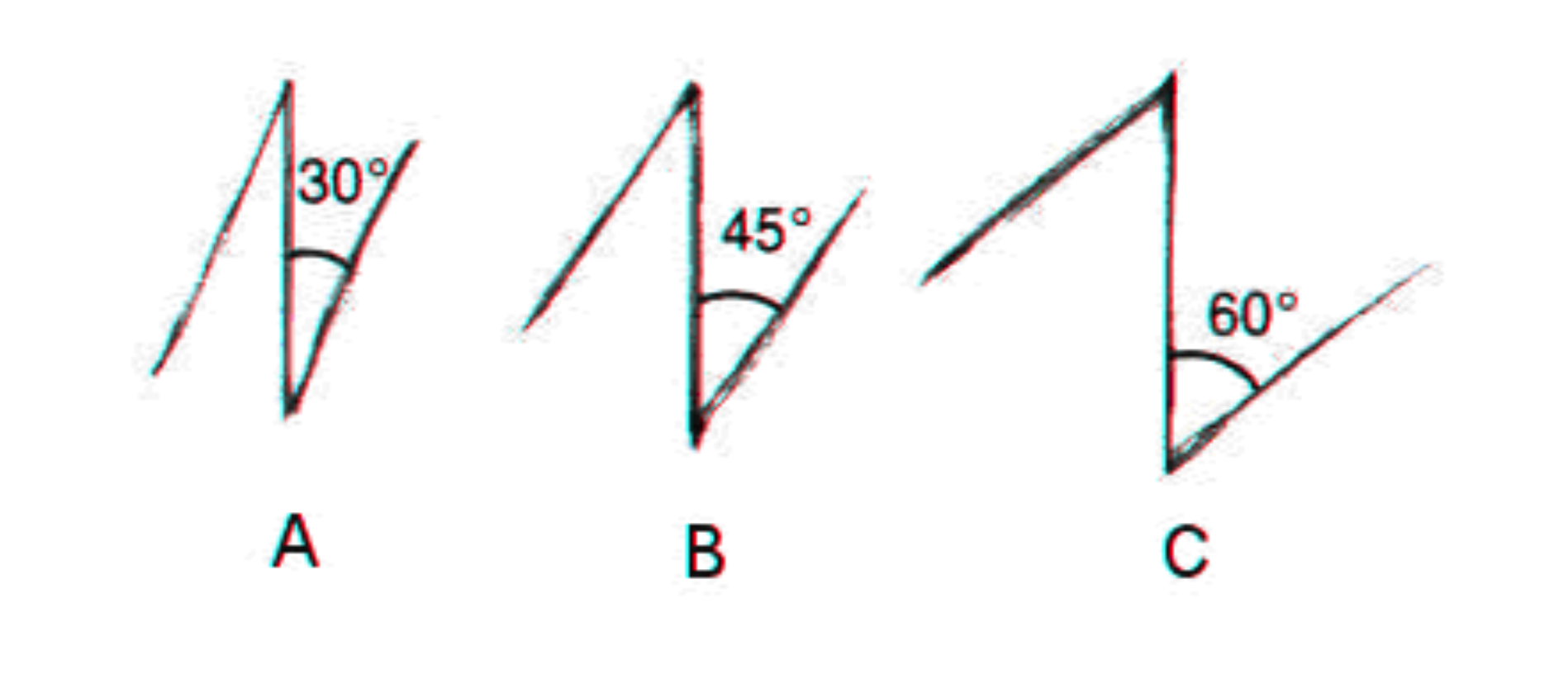   1. ~~A?~~ 2. ~~B?~~ 3. ~~C?~~ | 1. ~~25% (1 mark, max 1 mark)~~ 2. ~~50% (1 mark, max 1 mark)~~ 3. ~~75% (1 mark, max 1 mark)~~ |  |
| ~~13~~ | 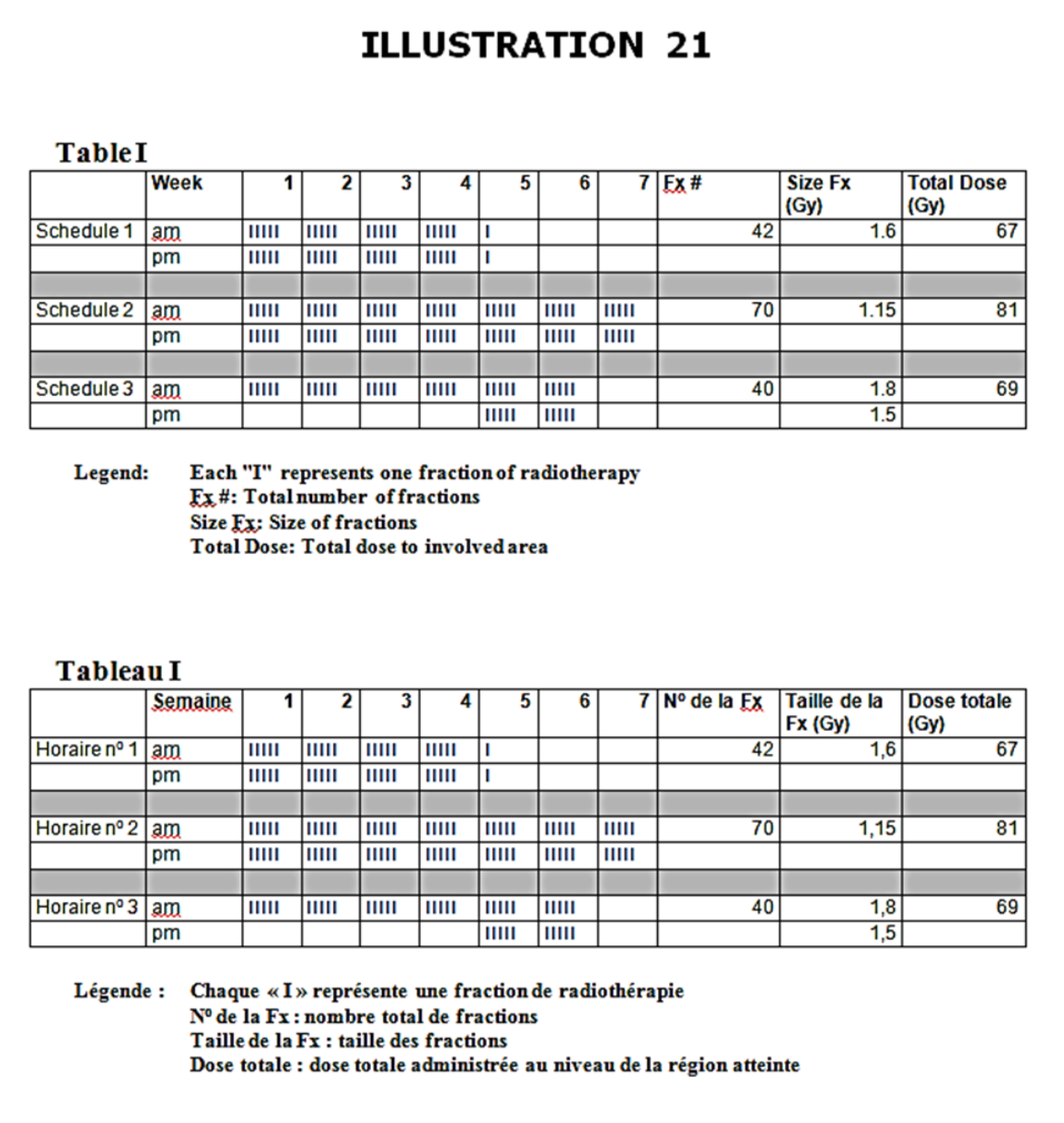  ~~See illustration.~~   1. ~~Name the types of radiation depicted in Table I.~~    a. ~~Schedule 1~~    b. ~~Schedule 2~~    c. ~~Schedule 3~~ 2. ~~What is the main radiobiologic rationale behind the type of radiation depicted in schedule 1?~~ 3. ~~What is the principal determinant of late radiation induced toxicity?~~ | ~~1 mark each, 5 marks max~~   1. a. ~~Accelerated~~    b. ~~Hyperfractionated~~    c. ~~Accelerated boost or concomitant boost~~ 2. ~~Reduce time for tumor to repopulate~~ 3. ~~Total dose~~ |  |
