## Supplement 5 for "Evaluating ChatGPT-4 in Otolaryngology–Head and Neck Surgery Board Examination using the CVSA Model"

| **Scenario no.** | **Scenario and questions** | **Model answer** | **GPT-4 answers (with lead in, used only in stem/prior to first question in set: “This is a question from an otolaryngology head and neck surgery licensing exam.”)** |
| --- | --- | --- | --- |
| 1 | A 6 year old boy is referred to you by his family physician. He has had 3 episodes of painful swelling of the right parotid gland over the past 18 months. Each episode lasted 5-7 days and has been accompanied by a low grade fever. Two of the episodes were managed with amoxicillin. One of the episodes was not treated with antibiotics and spontaneously resolved. There was complete resolution of symptoms between episodes. The child is otherwise healthy.   1. What is the most likely diagnosis? 2. List 4 treatment measures that are most commonly employed in the management of this condition. 3. What would you tell the parents about the long term clinical course of this disease? | 1. 1 mark, max 1 mark    a. Recurrent parotitis of childhood (must have entire term)    b. Also accept JRP (juvenile recurrent parotitis). 2. 0.5 marks for each, max 2 marks    a. Massage    b. Hydration    c. Analgesics    d. Antibiotics    e. Sialogogues    f. Heat (only 0.25 mark for this) 3. 1 mark each, max 1 mark    a. More recurrences    b. Probable remission as child enters adolescence    c. Sialoendoscopy | **Q1, A2**: The most likely diagnosis for this 6-year-old boy with recurrent episodes of painful swelling of the right parotid gland, accompanied by a low-grade fever, is recurrent parotitis. Recurrent parotitis is an inflammatory condition that affects the parotid gland and can be caused by various factors, such as infections, autoimmune diseases, or obstruction of the salivary ducts. In children, the most common cause is usually an obstruction or infection. Since the episodes resolved with or without antibiotic treatment and the child is otherwise healthy, this suggests an idiopathic etiology or a structural issue with the salivary ducts rather than an underlying systemic condition.  **Q2, A2**: The management of recurrent parotitis typically involves a combination of conservative measures and medical interventions to control symptoms, prevent complications, and address the underlying cause. Here are four commonly employed treatment measures:   1. Hydration and warm compresses: Encouraging the child to drink plenty of fluids and applying warm compresses over the affected area can help promote salivary flow and reduce glandular inflammation. 2. Sialogogues: Using sialogogues, such as sugar-free lemon drops or sour candies, can help stimulate salivary flow, which may help to clear any possible obstruction in the salivary ducts and reduce inflammation. 3. Antibiotics: If a bacterial infection is suspected or confirmed, a course of antibiotics, such as amoxicillin, may be prescribed to treat the infection. The choice of antibiotics should be guided by the local bacterial resistance patterns and the patient's medical history. 4. Pain relief and anti-inflammatory medications: Over-the-counter pain relievers and anti-inflammatory medications, such as ibuprofen or acetaminophen, can help to alleviate pain and reduce inflammation associated with recurrent parotitis.   In some cases, additional treatments may be necessary depending on the underlying cause of the recurrent parotitis. This may include surgical intervention to address any structural abnormalities or persistent obstructions, or the management of any underlying systemic conditions contributing to the recurrent episodes of parotitis.  **Q3, A2**: When discussing the long-term clinical course of recurrent parotitis with the parents, it's essential to provide them with an understanding of the condition and the potential outcomes. You could tell them the following:   1. In many cases, recurrent parotitis in children is self-limiting, meaning that it may resolve on its own over time. As the child grows, their salivary duct system may mature and develop, which can lead to a decrease in the frequency and severity of episodes. 2. Some children may continue to experience recurrent episodes throughout childhood, but the frequency and severity of these episodes may decrease over time. 3. In some cases, conservative and medical treatments may be sufficient to manage the symptoms, and the child may not require any further interventions. 4. In a minority of cases, more invasive procedures may be necessary to address the underlying cause, such as salivary duct abnormalities, persistent infections, or other underlying issues. 5. It is important to monitor the child's condition and follow up with the healthcare team regularly to assess their progress, adjust treatment plans if needed, and identify any potential complications or changes in their condition. 6. Overall, the prognosis for children with recurrent parotitis is generally good, and most children experience improvement or resolution of the condition as they grow older. However, the course of the disease can vary from one individual to another, and it is crucial to provide the child with appropriate care and support throughout their management. |
| 2 | You are performing an osteoplastic flap procedure using a bicoronal approach for drainage of mucoceles.   1. In which soft tissue planes will you elevate the bicoronal flap? 2. Once the frontal bone is exposed, list three techniques to identify margins of the frontal sinus prior to making the bony cuts for the osteoplastic flap. | 1. 1 mark each, 2 marks max    a. Subgaleal plane in the area between superior temporal lines    b. Superficial layer of deep temporal fascia overlying temporalis muscle    c. NO marks for subperiosteal plane 2. 1 mark each, 3 marks max    a. Plain x-ray used as template    b. Intraoperative image guidance/surgical navigation    c. Transillumination via frontal trephine | **Q1, A2**: During an osteoplastic flap procedure using a bicoronal approach for drainage of mucoceles, the bicoronal flap is elevated in the subgaleal and subperiosteal planes.   1. Subgaleal plane: The initial incision is made in the scalp, and the dissection proceeds through the skin and subcutaneous tissue. The galea aponeurotica is then identified, and the dissection continues in the subgaleal plane, which is the plane between the galea and the periosteum. The subgaleal dissection is performed anteriorly until reaching the supraorbital rims and posteriorly towards the occipital region, exposing the entire calvarial vault. 2. Subperiosteal plane: Once the bicoronal flap is elevated in the subgaleal plane, attention is turned to the periosteum. The periosteum is carefully incised, and a subperiosteal dissection is performed to expose the underlying bone. This dissection is performed carefully to preserve the integrity of the periosteum, which is essential for proper wound healing and prevention of complications like infection and hematoma.   By elevating the bicoronal flap in these two soft tissue planes, you gain access to the frontal bone, which can then be osteotomized, and the mucoceles can be drained effectively. After the procedure, the osteoplastic flap is replaced, and the layers are closed in a layered fashion.  **Q2, A2**: Once the frontal bone is exposed, it is crucial to accurately identify the margins of the frontal sinus before making the bony cuts for the osteoplastic flap. Here are three techniques to help identify the margins of the frontal sinus:   1. Digital palpation: One of the simplest methods to identify the sinus margins is by carefully palpating the supraorbital rims and the frontal bone. The frontal sinus is typically located above the orbits, and by palpating the area, you may feel the bony contours and transitions that indicate the sinus margins. 2. Preoperative imaging: Preoperative imaging studies, such as computed tomography (CT) scans, can provide valuable information about the size, shape, and location of the frontal sinus. It is essential to review these imaging studies before surgery to become familiar with the sinus's anatomy, which will help guide the dissection and identification of the sinus margins intraoperatively. 3. Intraoperative navigation: Intraoperative navigation systems, also known as image-guided surgery systems, can be utilized to assist in accurately identifying the frontal sinus margins. These systems utilize preoperative imaging data (e.g., CT scans) to provide real-time, three-dimensional guidance during surgery. By using a navigation probe, the surgeon can accurately identify the margins of the frontal sinus and avoid potential complications.   It is essential to accurately identify the frontal sinus margins to prevent injury to vital structures, such as the dura mater and the frontal lobe, and to ensure complete drainage and proper reconstruction of the osteoplastic flap. |
| 3 | List the three most common complications associated with placement of tympanostomy tubes in children. | 1 mark each, 3 marks max   a. Otorrhea b. Persistent perforation c. Granulation tissue formation | **A2**: Tympanostomy tubes, also known as ventilation tubes or pressure equalization (PE) tubes, are commonly placed in children to treat recurrent ear infections or persistent middle ear fluid. Though the procedure is generally safe, there are potential complications. The three most common complications associated with placement of tympanostomy tubes in children are:  Otorrhea: This refers to discharge or drainage from the ear, which can occur due to infection or inflammation. It is the most common complication and can usually be managed with antibiotic ear drops.  Persistent tympanic membrane perforation: Sometimes, the tympanic membrane (eardrum) fails to heal properly after the tube is removed or falls out, resulting in a persistent hole. This can lead to hearing loss, recurrent infections, or the need for further surgical intervention to repair the perforation.  Tube blockage or extrusion: The tubes can become obstructed by debris, blood, or infection, which may reduce their effectiveness. Alternatively, the tubes can be pushed out of the tympanic membrane prematurely, either due to the body's natural healing process or because of infection. Both situations may require additional treatment or surgical intervention. |
| 4 | Name 4 local oncologic contraindications to supra-cricoid laryngectomy. | 1 mark each, 4 marks max   a. Fixation of the arytenoid cartilage secondary to cricoarytenoid joint fixation b. Subglottic extension to the level of the cricoid or direct invasion of the cricoid    i. Paraglottic extension c. Invasion of the posterior commissure (interarytenoid area) d. Extension of the outer perichondrium of the thyroid cartilage or extralaryngeal spread | **A2**: Supra-cricoid laryngectomy is a surgical procedure that aims to preserve the patient's voice and swallowing function while removing a malignant tumor from the larynx. However, there are certain local oncologic contraindications that may preclude a patient from undergoing this procedure. Four local contraindications to supra-cricoid laryngectomy include:   1. Involvement of the pre-epiglottic space: If the tumor has invaded the pre-epiglottic space, supra-cricoid laryngectomy may not achieve adequate tumor clearance, and a more extensive surgical approach might be needed. 2. Involvement of the cricoid cartilage: The cricoid cartilage is an essential structure for the reconstruction of the larynx during supra-cricoid laryngectomy. If the tumor has infiltrated the cricoid cartilage, this surgery may not be feasible as it would compromise the functional outcome. 3. Thyroid cartilage invasion: If the tumor has extensively invaded the thyroid cartilage, supra-cricoid laryngectomy may not be suitable due to the risk of incomplete tumor resection and the potential impact on postoperative function. 4. Extension into the base of the tongue: Supra-cricoid laryngectomy might not provide adequate tumor clearance if the tumor has extended superiorly into the base of the tongue, necessitating alternative surgical approaches or other treatments.   The decision to perform a supra-cricoid laryngectomy is ultimately based on a thorough evaluation of the patient's tumor characteristics, overall health, and the potential risks and benefits of the procedure. |
| 5 | A patient presents with an anterior glottic web. Name 5 treatment options, used alone or in combination, for the management of this lesion. | 0.5 marks each, 2.5 marks max   a. Observation b. CO_2_ laser c. Cold dissection d. Mitomycin C e. Laryngofissure with keel insertion f. Steroid injection g. Endoscopic keel | **A2**: An anterior glottic web is a fibrous or membranous tissue that connects the anterior part of the vocal cords. This condition can result in voice disorders, airway obstruction, and other complications. Several treatment options can be used alone or in combination to manage an anterior glottic web, including:   1. Endoscopic lysis: This involves using an endoscope and microsurgical instruments to cut and remove the web. It is a minimally invasive procedure and can be performed under general anesthesia. 2. Microflap excision: In this technique, a small flap of mucosa is elevated from the vocal cord, and the web is carefully dissected and removed. The flap is then repositioned and sutured in place to cover the defect. 3. CO2 laser ablation: A CO2 laser can be used to vaporize the web tissue precisely and with minimal damage to the surrounding vocal cord tissue. This technique can be combined with other surgical procedures for optimal results. 4. Keel placement: Following the removal of the web, a temporary stent, or keel, can be placed between the vocal cords to prevent the reformation of the web during the healing process. The keel is usually removed after a few weeks. 5. Laryngotracheal reconstruction: In severe cases, where the web is causing significant airway obstruction, a laryngotracheal reconstruction may be necessary. This procedure involves surgically enlarging the airway and reconstructing the vocal cords to restore normal function.   Note that treatment decisions should be tailored to the individual patient and the severity of the web. A multidisciplinary team, including an otolaryngologist, speech-language pathologist, and other specialists, should be involved in the management and rehabilitation process to ensure optimal outcomes. |
| 6 | Name 3 techniques used to prevent or cure pharyngoesophageal spasm after total laryngectomy. | 1 mark each, 3 marks max   a. Cricopharyngeal myotomy b. Pharyngeal plexus neurectomy c. Botox injection | **A2**: Pharyngoesophageal spasm after total laryngectomy can lead to difficulty with swallowing and other complications. Various techniques can be employed to prevent or cure this issue. Here are three commonly used techniques:   1. Botulinum Toxin (Botox) Injection: Botox injections can be administered to the pharyngoesophageal segment to relax the muscle and reduce spasms. The effect of the injection is temporary, usually lasting for a few months, and may need to be repeated as necessary. 2. Pharyngeal Myotomy: This is a surgical procedure that involves cutting the muscle fibers of the pharyngoesophageal segment to reduce spasms and improve swallowing function. It is often performed when other conservative treatments have failed to provide relief. 3. Balloon Dilation: This technique involves the use of a balloon catheter, which is inserted into the pharyngoesophageal segment and gradually inflated to dilate the area and alleviate the spasm. This method can be repeated if necessary and is often used in conjunction with other treatments like Botox injections or pharyngeal myotomy.   These techniques can provide relief to patients experiencing pharyngoesophageal spasms after total laryngectomy. The choice of technique may depend on the severity of the spasms, patient preferences, and the surgeon's expertise. It is essential to evaluate the patient carefully and develop an individualized treatment plan for optimal outcomes. |
| 7 | What is the main arterial supply to each of the following flaps once they are elevated?   1. Pectoralis major flap 2. Latissimus dorsi flap 3. Lower island trapezius flap 4. Paramedian forehead flap 5. Deltopectoral flap 6. Fibular free flap 7. Iliac osteomyocutaneous free flap | 0.5 marks each, 3.5 marks max   1. Pectoral branch of thoracoacromial    a. Lateral thoracic (0.25 marks only) 2. Thoracodorsal 3. Transverse cervical    a. Dorsal scapular (0.25 marks only) 4. Supratrochlear 5. Perforator intercostal of the internal mammary 6. Peroneal 7. Deep circumflex iliac | **Q1, A2**: The main arterial supply to the pectoralis major flap, once elevated, is the thoracoacromial artery. This artery is a branch of the axillary artery, and it provides blood supply to the pectoralis major muscle, which is the primary component of the flap. Additionally, perforating branches from the internal mammary artery can also contribute to the vascularization of the flap.  **Q2, A2**: The main arterial supply to the latissimus dorsi flap, once elevated, is the thoracodorsal artery. The thoracodorsal artery is a branch of the subscapular artery, which itself arises from the axillary artery. The thoracodorsal artery provides blood supply to the latissimus dorsi muscle, the primary component of the flap.  **Q3, A2**: The main arterial supply to the lower island trapezius flap, once elevated, is the transverse cervical artery. Specifically, the descending branch of the transverse cervical artery supplies the lower part of the trapezius muscle. The transverse cervical artery arises from the thyrocervical trunk, which is a branch of the subclavian artery. This vascular supply enables the lower island trapezius flap to be used in reconstructive procedures.  **Q4, A2**: The main arterial supply to the paramedian forehead flap, once elevated, is the supratrochlear artery. The supratrochlear artery is a branch of the ophthalmic artery, which itself is a branch of the internal carotid artery. The supratrochlear artery supplies blood to the forehead and is responsible for providing vascularization to the skin and subcutaneous tissue of the paramedian forehead flap.  **Q5, A2**: The main arterial supply to the deltopectoral flap, once elevated, is the internal mammary artery, also known as the internal thoracic artery. The internal mammary artery is a branch of the subclavian artery. It runs down the chest wall and provides blood supply to the deltopectoral groove, which is the primary component of the flap. Additionally, perforating branches from the thoracoacromial artery can contribute to the vascularization of the flap.  **Q6, A2**: The main arterial supply to the fibular free flap, once elevated, is the peroneal artery, also known as the fibular artery. The peroneal artery is a branch of the tibial-fibular trunk, which itself arises from the popliteal artery. The peroneal artery provides blood supply to the fibula bone, as well as the surrounding soft tissue and skin, making it the primary vascular source for the fibular free flap used in reconstructive procedures.  **Q7, A2**: The main arterial supply to the iliac osteomyocutaneous free flap, once elevated, is the deep circumflex iliac artery (DCIA). The deep circumflex iliac artery is a branch of the external iliac artery, which originates from the common iliac artery. The DCIA supplies blood to the iliac crest bone, the overlying skin, and the accompanying soft tissue, making it the primary vascular source for the iliac osteomyocutaneous free flap used in reconstructive procedures. |
| 8 | A 49 year old patient with T3N2b squamous cell carcinoma of the hypopharynx is scheduled to receive combined chemotherapy and radiation. What is the most important potential long-term sequelae of this treatment that must be disclosed to the patient? | 1 mark, 1 mark max   a. Permanent dysphagia/need for tube feeding    i. Airway obstruction or xerostomia (0.5 mark only) | **A2**: The most important potential long-term sequelae of combined chemotherapy and radiation for T3N2b squamous cell carcinoma of the hypopharynx that must be disclosed to the patient include the risk of dysphagia, xerostomia, and radiation-induced malignancy.  Dysphagia, or difficulty swallowing, is a common side effect of radiation therapy to the hypopharynx, and can result in the need for a feeding tube or prolonged difficulty with eating and drinking.  Xerostomia, or dry mouth, is another common side effect of radiation therapy to the head and neck, and can result in difficulty with speaking, swallowing, and increased risk of dental caries and oral infections.  Lastly, radiation-induced malignancy, although rare, can occur years after radiation therapy and must be discussed with the patient. This risk increases with higher doses of radiation and longer follow-up periods.  It is important to note that these potential side effects should be discussed with the patient in the context of the potential benefits of treatment and the overall prognosis of the disease. |
| 9 | List four genera of fungi that cause mucormycosis. | 0.5 mark each, 2 marks max   a. Mucor b. Rhizomucor c. Rhizopus d. Absidia e. Apophysomyces | **A2**: Mucormycosis, also known as zygomycosis, is a severe and potentially life-threatening infection caused by fungi belonging to the order Mucorales. The following are four genera of fungi that commonly cause mucormycosis:   1. Rhizopus 2. Mucor 3. Lichtheimia (formerly Absidia) 4. Cunninghamella   These fungi are opportunistic pathogens and typically cause infection in immunocompromised individuals, such as those with poorly controlled diabetes, organ transplant recipients, or patients undergoing chemotherapy. Mucormycosis can manifest in various clinical forms, including rhinocerebral, pulmonary, cutaneous, gastrointestinal, and disseminated infections. Early diagnosis and aggressive treatment, which may involve antifungal therapy and surgical intervention, are crucial to improve patient outcomes. |
| 10 | 1. List 2 functions of the superior laryngeal nerve. 2. Where do the recurrent laryngeal nerve and the internal branch of the superior laryngeal nerve enter the larynx respectively? | 1. 0.5 marks each, 1 mark max    a. Internal branch - sensory to the mucosa of the larynx superior to the true vocal cord       i. *need to be specific as to where it provides sensation    b. External branch - motor to the cricothyroid muscle 2. 0.5 marks each, 1 mark max    a. Recurrent nerve enters posterior to the cricothyroid joint    b. Internal branch of the superior laryngeal nerve pierces the thyrohyoid membrane along with the superior laryngeal artery       i. *detail about artery as mentioned above not required to receive point | **Q1, A2**: The superior laryngeal nerve is a branch of the vagus nerve (cranial nerve X) and plays an important role in the functioning of the larynx. It has two primary functions:   1. Sensory function: The internal branch of the superior laryngeal nerve provides sensory innervation to the laryngeal mucosa above the vocal cords, including the epiglottis, aryepiglottic folds, and arytenoid cartilages. This sensory function helps in the protective reflexes such as coughing and swallowing, preventing aspiration of foreign materials into the airway. 2. Motor function: The external branch of the superior laryngeal nerve supplies motor innervation to the cricothyroid muscle. The cricothyroid muscle is responsible for tensing and elongating the vocal cords, which results in an increase in pitch during phonation.   **Q2, A2**: The recurrent laryngeal nerve and the internal branch of the superior laryngeal nerve enter the larynx at different locations:   1. Recurrent laryngeal nerve: The recurrent laryngeal nerve enters the larynx by passing underneath the lower border of the inferior constrictor muscle, posterior to the cricothyroid joint. It then ascends in the tracheoesophageal groove to provide motor innervation to all intrinsic laryngeal muscles, except the cricothyroid muscle, and sensory innervation to the laryngeal mucosa below the vocal cords. 2. Internal branch of the superior laryngeal nerve: The internal branch of the superior laryngeal nerve enters the larynx through the thyrohyoid membrane, which is situated between the thyroid cartilage and the hyoid bone. It provides sensory innervation to the laryngeal mucosa above the vocal cords, including the epiglottis, aryepiglottic folds, and arytenoid cartilages. |
| 11 | List all of the neural and vascular structures that pass through the sphenopalatine foramen. | 1 mark each, 4 marks max   a. Sphenopalatine artery b. Sphenopalatine vein c. Nasopalatine nerve d. Posterosuperior lateral nasal nerve | **A2**: The sphenopalatine foramen is a small opening in the skull that connects the pterygopalatine fossa to the nasal cavity. It allows the passage of specific neural and vascular structures. The main structures that pass through the sphenopalatine foramen include:   1. Sphenopalatine artery: This artery is a branch of the maxillary artery and supplies the nasal cavity, nasopharynx, and the adjoining structures. 2. Nasopalatine nerve: This nerve is a branch of the pterygopalatine ganglion, which is itself a branch of the maxillary nerve (CN V2). The nasopalatine nerve provides sensory innervation to the nasal septum and the anterior part of the hard palate. 3. Posterior superior nasal nerves: These nerves are also branches of the pterygopalatine ganglion, and they provide sensory innervation to the posterior and superior parts of the nasal cavity. 4. In addition to these primary structures, the sphenopalatine foramen may also transmit small branches of the pterygopalatine ganglion that contribute to the innervation of the nasal mucosa and some blood vessels that supply the surrounding structures. |
| ~~12~~ | ~~Theoretically, by what percentage will the Z-plasties shown in this referenced illustration increase scar length?~~  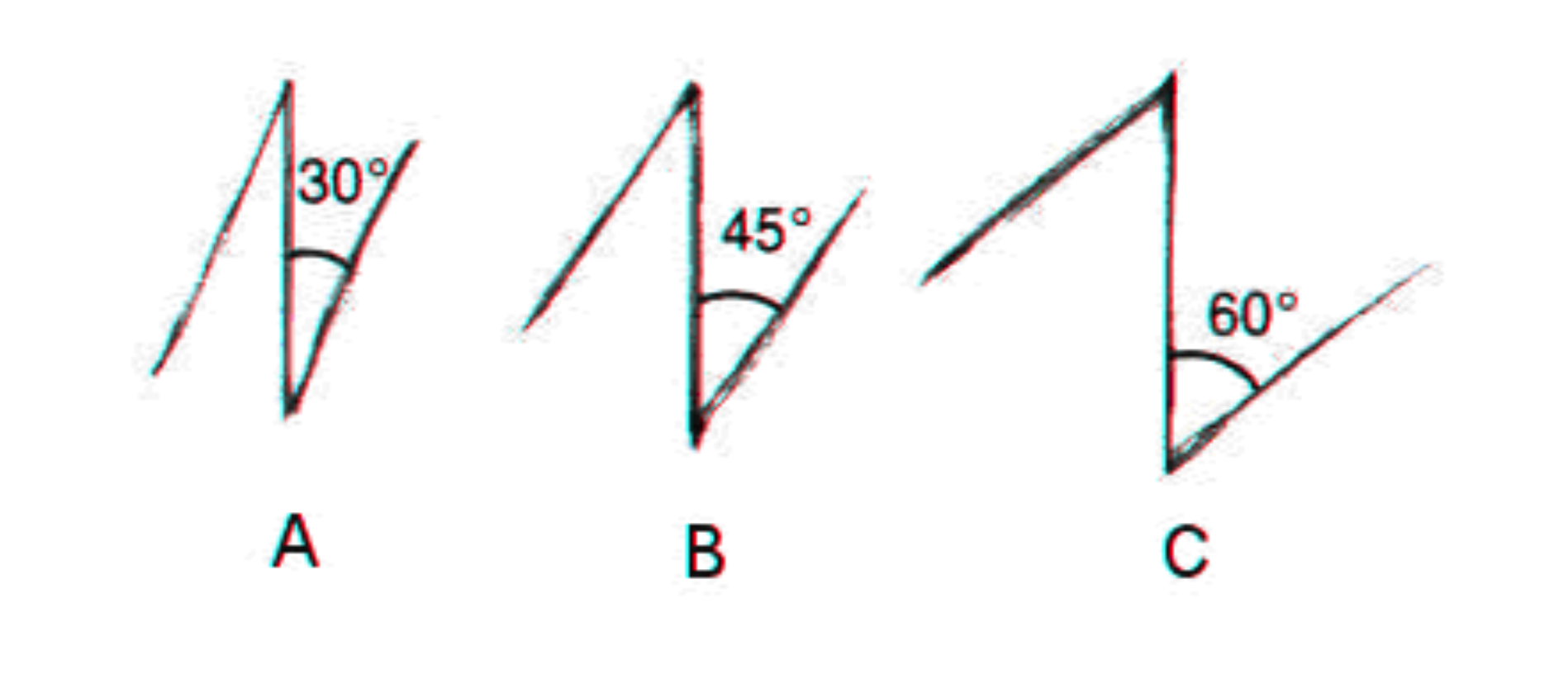   1. ~~A?~~ 2. ~~B?~~ 3. ~~C?~~ | 1. ~~25% (1 mark, max 1 mark)~~ 2. ~~50% (1 mark, max 1 mark)~~ 3. ~~75% (1 mark, max 1 mark)~~ |  |
| ~~13~~ | 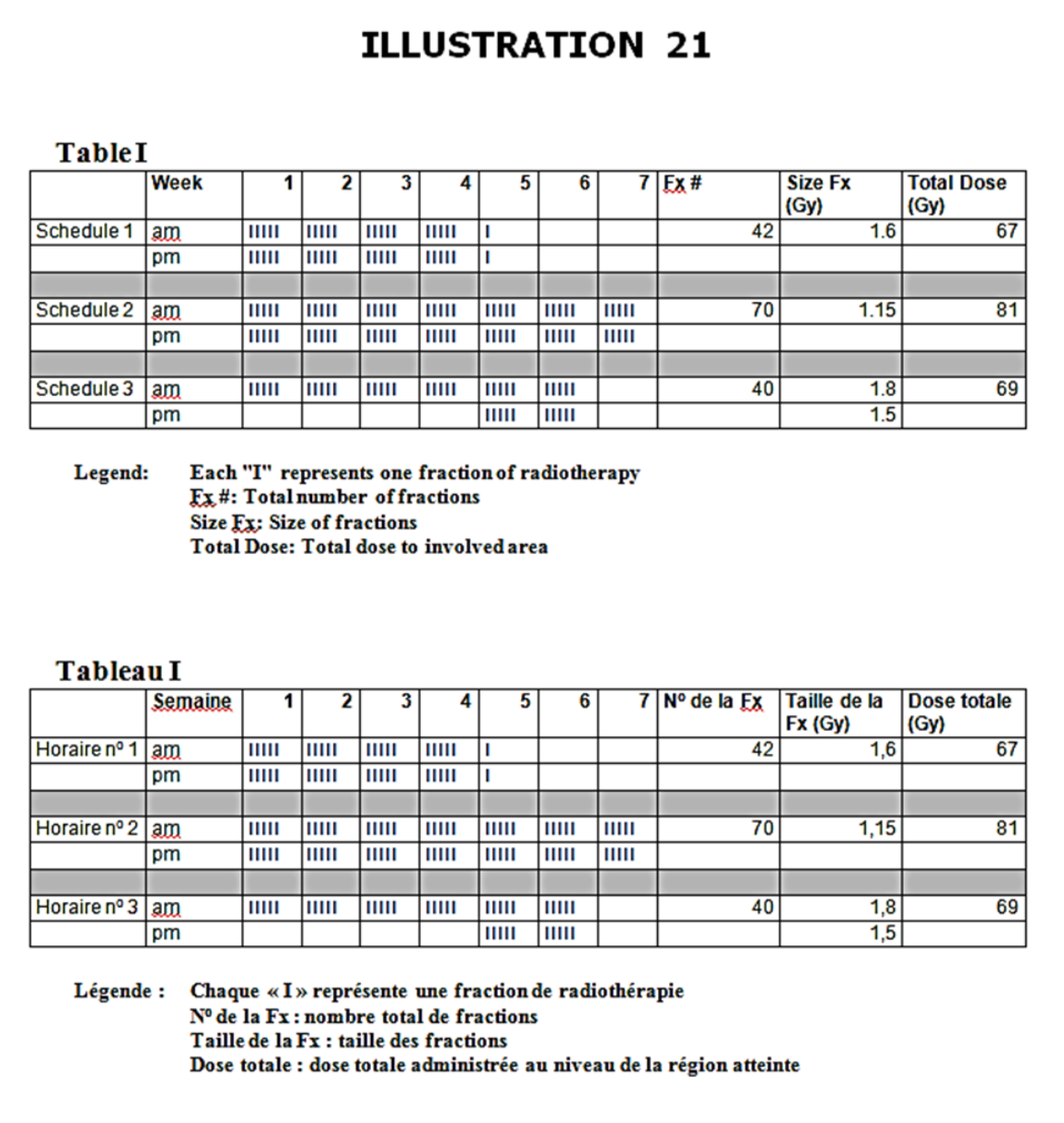  ~~See illustration.~~   1. ~~Name the types of radiation depicted in Table I.~~    a. ~~Schedule 1~~    b. ~~Schedule 2~~    c. ~~Schedule 3~~ 2. ~~What is the main radiobiologic rationale behind the type of radiation depicted in schedule 1?~~ 3. ~~What is the principal determinant of late radiation induced toxicity?~~ | ~~1 mark each, 5 marks max~~   1. a. ~~Accelerated~~    b. ~~Hyperfractionated~~    c. ~~Accelerated boost or concomitant boost~~ 2. ~~Reduce time for tumor to repopulate~~ 3. ~~Total dose~~ |  |
